# Operational response simulation tool for epidemics within refugee and IDP settlements

**DOI:** 10.1101/2021.01.27.21250611

**Authors:** Joseph Aylett-Bullock, Carolina Cuesta-Lazaro, Arnau Quera-Bofarull, Anjali Katta, Katherine Hoffmann Pham, Benjamin Hoover, Hendrik Strobelt, Rebeca Moreno Jimenez, Aidan Sedgewick, Egmond Samir Evers, David Kennedy, Sandra Harlass, Allen Gidraf Kahindo Maina, Ahmad Hussien, Miguel Luengo-Oroz

## Abstract

The spread of infectious diseases such as COVID-19 presents many challenges to healthcare systems and infrastructures across the world, exacerbating inequalities and leaving the world’s most vulnerable populations most affected. Given their density and available infrastructure, refugee and internally displaced person (IDP) settlements can be particularly susceptible to disease spread. Non-pharmaceutical public health interventions can be used to mitigate transmission, and modeling efforts can provide crucial insights on the potential effectiveness of such interventions to help inform decision making processes. In this paper we present an agent-based modeling approach to simulating the spread of disease in refugee and IDP settlements. The model, based on the JUNE open-source framework, is informed by data on geography, demographics, comorbidities, physical infrastructure and other parameters obtained from real-world observations and previous literature. Furthermore, we present a visual analytics tool which allows decision makers to distill insights by comparing the results of different simulations and scenarios. Through simulating their effects on the epidemiological development of COVID-19, we evaluate several public health interventions ranging from increasing mask wearing compliance to the reopening of learning institutions. The development and testing of this approach focuses on the Cox’s Bazar refugee settlement in Bangladesh, although our model is designed to be generalizable to other informal settings.

## 1 Introduction

The spread of COVID-19 across the world presents many challenges to healthcare systems and infrastructures, exacerbating inequalities and leaving the world’s most vulnerable populations most affected. Refugee and internally displaced persons (IDPs) settlements, especially those which have been rapidly created in response to sudden crises, often suffer from overcrowding and insufficient sanitation facilities. Furthermore, some of these settlements are located in regions with frequent natural disasters such as flooding and earthquakes or conflict zones. Given these conditions, disease spread in settlements has previously been shown to be rapid [1]. The COVID-19 pandemic presents significant threats to people living in these settlements, and the provision of detailed information on potential mitigation strategies is of vital importance. In this paper, we present a simulation tool to support decision making and advocacy by simulating the potential effectiveness of operational interventions in refugee and IDP settlements.

Specifically, we take an agent-based modeling (ABM) approach to understand the impact of public health interventions on limiting disease spread in settlements. Operational interventions (particularly non-pharmaceutical interventions) can take a variety of forms, from alternative care delivery mechanisms to behavioral interventions such as mask wearing and physical distancing. In settlements, some of the most frequent interventions for mitigating disease spread are not feasible due to the complex environments and difficult conditions in which Persons of Concern (PoCs) live.^1^ For example, a lack of Personal Protective Equipment (PPE) such as surgical masks could make well-established measures such as compulsory mask wearing challenging to implement. To overcome these difficulties, new operational interventions must also be devised and evaluated. By simulating the possible effects of such measures, we hope to provide teams on the ground with reliable insights for data-driven decision making and situational planning.

The development and testing of this approach focuses on the Cox’s Bazar refugee settlement in Bangladesh, which reported its first COVID-19 case in mid-May 2020 [3]. In particular, we analyze operational interventions by modeling the interactions between residents of the Kutupalong-Batukhali Expansion Site. With over 900,000 Rohingya PoCs, Cox’s Bazar contains one of the largest refugee settlements in the world [4]. Its inhabitants are primarily Rohingya people, a stateless Muslim minority who have continuously fled targeted violence, discrimination, and human rights violations in Myanmar [5]. A number of risk factors make the settlement vulnerable to epidemic outbreaks. The settlement has high rates of global acute malnutrition and other comorbidities such as respiratory illnesses, which could lead to lower general immunity among camp residents [6]. Population density is exceptionally high and many facilities are communal, increasing the risk of person-to-person transmission. Finally, limited access to sources of information such as the internet, as well as low levels of literacy, make public health campaigns challenging.

Given the significant risk of disease spread in a settlement as dense as the Cox’s Bazar settlement, UNHCR and World Health Organization (WHO) teams responded rapidly to the COVID-19 pandemic, initiating preventative activities two months before the first case was confirmed [7, 8, 9]. Testing and case reporting are challenging in the settlement setting and the available data is limited. For this reason, we take a scenario-based approach focusing on simulating the relative efficacies of potential interventions, as opposed to attempting to predict highly accurate numbers for infections, hospitalization and fatality rates which would require more complete data for model fitting. As a result of this design choice, detailed COVID-19 case data is not required for our modeling; instead, we rely on a set of clearly recorded assumptions on interaction and transmission probabilities which can be varied in sensitivity analyses.

The structure and functionality of our model have been adapted from JUNE [10], a generalizable ABM framework for modeling the movement and interactions of people at the individual level, which was first used to model the spread of COVID-19 in England. Our methodology has been designed to apply not only to the current COVID-19 situation, but also to generalize to situation planning in future disease outbreaks in similar geographies. Our modeling process consists of four stages: i) building a ‘digital twin’ of the community of interest; ii) understanding and simulating the possible movements and interactions of the community’s residents; iii) implementing operational interventions to simulate their effects on the spread of disease; and iv) communicating findings to decision makers and experts in the field. This final step is equally as important as the others since if results cannot be effectively communicated, then valuable insights from the model will not be useful.

Agent-based models (ABMs) often require significant data inputs and computational resources which can create a barrier to their use in data- and resource-sparse environments. However, many humanitarian settlements throughout the world follow standardised reporting guidelines, and therefore may even have more detailed information available on demographics, geography, and the locations of amenities than in other national-level settings. Much of this data is regularly updated and made freely available by United Nations (UN) agencies and Non-Governmental Organisations (NGOs) operating in the settlements. To facilitate the generalizability of our approach to other settlements, we base our model on publicly-available data accessed from the Humanitarian Data Exchange (HDX) [11], as well as other data sources available upon request such as the UNHCR Microdata Library [12] and WHO Early Warning, Alert and Response System (EWARS) data [13]. The simulations presented in this report can each be run on a single processing unit of a laptop in approximately two hours^2^ for a settlement the size of that in Cox’s Bazar. This means that the model could be deployed even in settings where computing resources may be limited.

The core strengths of our approach are that (i) we develop a framework for simulating epidemics in complex refugee and IDP settings that takes into account highly detailed data on geography, population structure and behavior, facilities and potential mixing points; (ii) we can implement operational interventions as changes in parameters such as movement patterns, social behaviours or contact intensity, which makes it possible to evaluate a wide range of policy options - even geographically heterogeneous - without fundamentally altering the model structure; (iii) we account for detailed modelling of health trajectories including impacts of different comorbidities, and (iv) we propose a visual analytics framework that allows to distil insights from the simulations of particular operational interventions which is designed to be used by public health experts and decision makers.

Throughout this paper, we present our modeling approach, how our results are derived, and the parameters used such that this work can be reproduced and adapted to other settings. In Section 4 we describe how the ‘digital twin’ of the camp is created, drawing primarily on open-source census datasets to create a virtual population endowed with demographic attributes. Section 5 focuses specifically on how individuals move and interact in our model, based on reporting and insights from teams operating in the Cox’s Bazar settlement as well as epidemiological considerations of COVID-19 previously implemented in JUNE [10]. We discuss a variety of operational interventions implemented in our simulations and present the findings in Section 6. Finally, to complement our simulations, we have developed a tool to visualize and dynamically interact with results to aid decision-makers with the interpretation of our findings, which we describe in Section 7.

## 2 Related work

### 2.1 Epidemiological Modeling

Modeling approaches to infectious diseases span a broad range of techniques. Some of the most common methodologies are differential equation-based compartmental models such as Susceptible-Infectious-Recovered (SIR) and Susceptible-Exposed-Infectious-Recovered (SEIR) models [14]. These approaches are useful for gaining high-level insights; predictions are generally based on aggregate data and can allow for flexibility and transferability between regions. While some compartmental approaches have been developed to offer geographically dis-aggregated modeling, the level of granularity allowed is generally low and populations are typically considered to be homogeneously distributed within each geographical region.

As an alternative, agent-based models are often chosen due to their ability to capture the heterogeneous distribution of a population, both geographically and demographically, as well as differences in behavioral patterns [15, 16, 17]. ABMs offer the possibility to model group-level dynamics and social mixing down to the individual level, where disease spread is usually modeled through agent-to-agent contact (see for example [18, 19]). This level of granularity requires more detailed data inputs and increased computing power relative to compartmental approaches. However, with mass data collection efforts by governments and institutions becoming the norm and computational hardware and software allowing for more complex models, ABMs are becoming increasingly viable options for disease modeling.

Given the granular nature of these models and the ability to finely adjust the behaviors of the simulated population, ABMs are an powerful class of models for evaluating operational interventions for disease mitigation and can complement their compartmental counterparts. In practice, applications of ABMs to epidemiological settings can be quite diverse. A literature review by Hunter *et al*. [20] highlights a number of important axes of variation between models, including: whether disease transmission is person-to-person, vector-borne, or by ingestion of contaminated food or water; how transmission patterns incorporate population density and/or social network structure; and the actions that are available to agents when responding to the threat of infection.

### 2.2 ABMs in low-resource settings

There have been several studies that use ABMs for modeling infectious disease in low-resource settings. For example, previous research has modeled the spread of cholera in the Dadaab refugee camp in Kenya and a settlement in Ghana, as well as the spread of Ebola in Liberia [21, 22, 23]. More generally, Anderson *et al*. [24] construct a generic model of refugee camps to simulate potential health risks, allowing for the presence of multiple actors in the response (e.g., media, government institutions, and camp organizations) and for different policy interventions (such as providing medical supplies, food, and water). Practically speaking, our work is most similar to Crooks and Hailegiorgis [21] and Augustijn-Beckers *et al*. [22], whose models of cholera spread also incorporate detailed information such as the geographic structure of settlements, heterogeneous agents embedded in households, and the movement of agents to undertake routine daily activities. However, like the Ebola model of Merler *et al*. [23], our primary infection mechanism is person-to-person transmission through the simultaneous physical presence of agents in a location, and we simulate policy interventions to mitigate the spread of disease.

### 2.3 Models of COVID-19 in settlements

More recently, a number of compartmental and agent-based models have been developed to simulate the spread of COVID-19 in refugee and IDP settlements specifically. In the context of the Cox’s Bazar settlement, Truelove *et al*. [25] present a compartmental modeling approach simulating the spread of COVID-19 in the Kutupalong-Batukhali Expansion Site with a focus on predicting infection, hospitalization, and mortality statistics for operational planning based on different transmission scenarios defined by their reproduction number. Gilman *et al*. [26] take an agent-based approach to modeling the spread of the disease in the Moria camp in Greece and assess several possible intervention measures.

While our work shares many of the fundamental features of these approaches, we build on this and prior research on ABMs in low-resource settings based on three pillars. First, we provide a framework for modeling more complex settings with a larger population and a larger number of potential mixing points than the existing COVID-19 models described above. While our model structure is generally applicable and not limited to a given disease or setting (in a similar spirit to Anderson *et al*. [24]), our model allows the user to incorporate detailed population-specific data, interactions, and geographical structures that form the strengths of many of the ABMs described above.

Second, our model is designed to facilitate the simulation and comparison of different operational interventions. Practically speaking, these interventions are modeled as changes in agent behavior (i.e. movement patterns) and contact intensity rather than model *structure*, which makes implementing interventions a generic process. This means that we are not restricted to specific types of interventions, but rather can essentially model any intervention that operates through changing movement or contact intensity.

Third, we enhance our ABM with a methodology that accounts for comorbidities and allows for the incorporation of data on differences in access to healthcare systems across populations. Specifically, we explicitly incorporate age- and sex-specific comorbidity distributions and their influence on disease severity, which allows us to tailor hospitalization and mortality rates to context-specific population structures.

## 3 Context - Cox’s Bazar Refugee Settlement

We focus on the Cox’s Bazar refugee settlement in Bangladesh, which is one of the largest and most congested settlements in the world. Over 742,000 refugees arrived in the Cox’s Bazar settlement after an outbreak of violence in Myanmar’s Rakhine State in 2017, forming the largest historical influx of Rohingya into Bangladesh [27]. The Kutupalong-Batukhali Expansion Site (shown in Figure 1) refers to the set of camps located in Ukhia province excluding the original Kutupalong Refugee Camp, and is the most densely populated and most interconnected site in the settlement. The expansion site contains an estimated 583,000 [28] inhabitants, the majority of whom arrived during the influx in 2017 [29]. However, the expansion site spans only 13 square kilometers, with as few as 8 square meters of space available per person in some areas [5].

**Figure 1:**
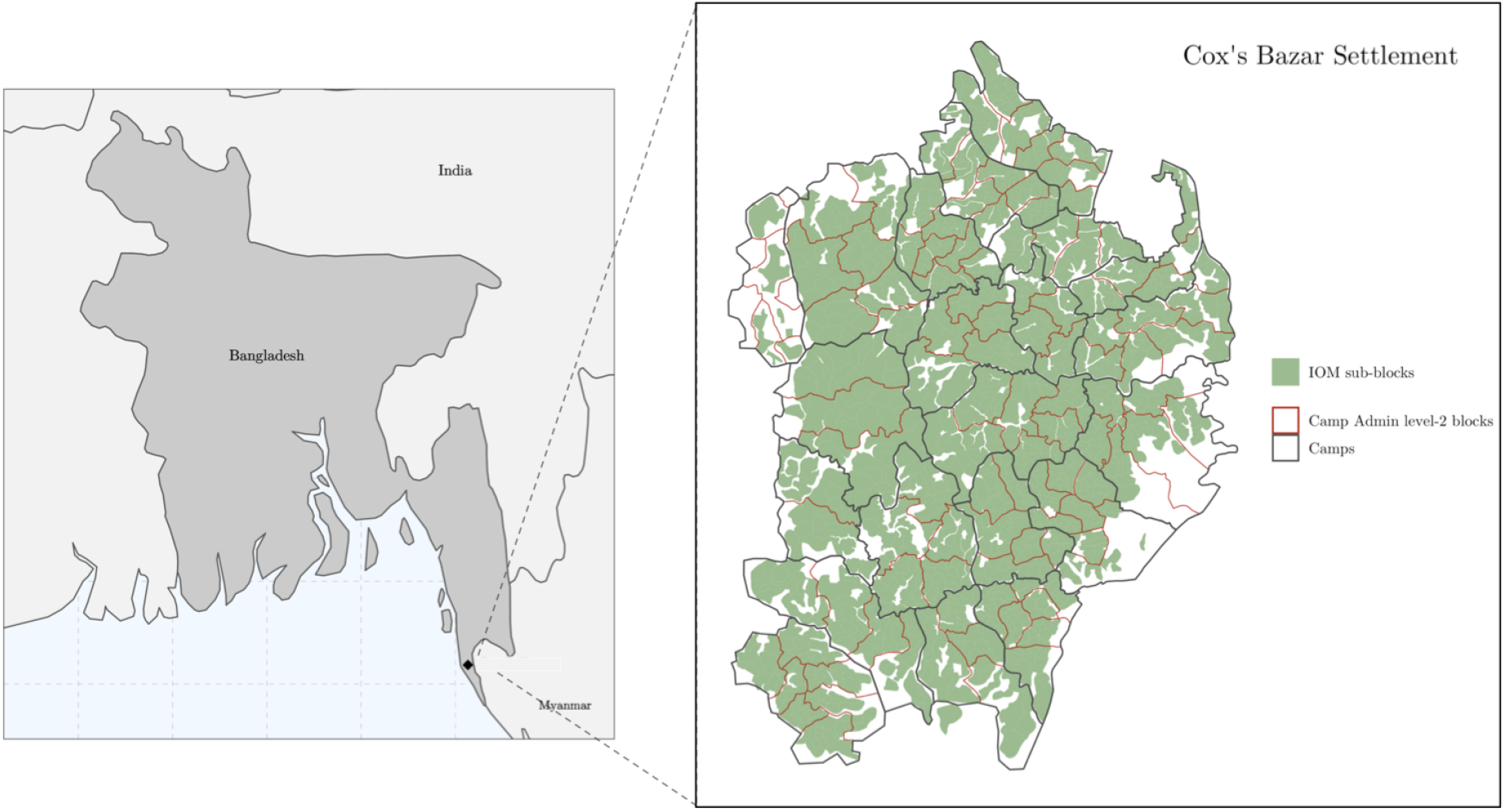
Left: Map of Bangladesh showing location of the Cox’s Bazar settlement. Right: Map of the modelled expansion site: black boundaries denote the camps, red boundaries denote camp Admin level-2 blocks and green shaded areas denote the areas in which people reside in sub-blocks as defined by IOM.

The majority of PoCs seeking refuge in Bangladesh are women and children, with more than 40% of PoCs under the age of 12 [27]. The PoCs in the Cox’s Bazar settlement are unable to formally participate in the economy of the host community and only have access to limited forms of livelihood. Consequently, “the refugee population remains 100% reliant on food assistance” [30]. In addition to food assistance, the United Nations Refugee Agency (UNHCR) and other humanitarian partners provide other necessary supplies (e.g., water, sanitation and hygiene (WASH) and non-food items (NFIs)) and facilitate activities such as informal education, livelihood programs, and community centers and safe spaces [31, 32].

A number of characteristics of the settlement make it vulnerable to the outbreak of a respiratory illness such as COVID-19. Many shelters are shared by multiple households and tend to be overcrowded [5]. PoCs also rely on publicly shared pumps, latrines, and other sanitation facilities making it harder to contain the potential spread of infectious diseases [33]. Poor nutrition combined with low general immunization coverage in the settlement makes PoCs vulnerable to communicable infections from both vaccine-preventable and water-borne diseases [6, 34, 35, 36]. Although hospitals and other health services are available to PoCs in the camp, resources, capacity, health personnel, and overall infrastructure are limited, making it difficult to treat a large-scale outbreak of a disease such as COVID-19. UNHCR, WHO and other humanitarian partners have worked quickly to put in place a variety of measures to address these challenges, such as the wide distribution of soap and then significant expansion of hospital capacity [37, 38, 39]. However, the exploration of additional strategies to mitigate the potential spread of the disease are imperative in this resource-constrained context.

## 4 Digital Twin

The first stage of our modeling process requires building a ‘digital twin’ of the settlement. This consists of defining the geographical structure of the model, building the virtual population and assigning demographic attributes, and constructing locations where individuals can interact with each other. This construct forms the basis for the environment in which the simulation can be run.

### 4.1 Geography

The geographical layout of a given settlement can vary significantly and impacts the distribution and density of its inhabitants. The JUNE model allows users to define three geographical levels in increasing order of granularity: regions, super areas, and areas [10].

There are multiple ways to divide the Cox’s Bazar settlement into these geographical levels by following different conventions. We make the decision based on readily available population and geographical data. Specifically, for ‘regions’ we select the camps which make up the Expansion Site to be the most aggregate level at which we model. These camps contain between 6,000 - 40,000 people each [28]. For the middle geographical layer (‘super areas’) we use the camp Admin level 2 blocks [40]; each camp contains 6-8 blocks. Finally, for the highest level of granularity (‘areas’), we use the sub-blocks as defined by the International Organization for Migration (IOM) [41].

Figure 1 shows the three different geographical layers used in our model of the refugee settlement in Cox’s Bazar. Using shapefiles provided by UNHCR for both the camps and the admin level 2 blocks [40], as well as shapefiles from IOM for the sub-blocks [41], we encode the geographical hierarchy by assigning camp Admin level 2 blocks to the camps with which they overlap the most, and by assigning the IOM sub-blocks to the the Admin level 2 blocks with which they overlap the most.

### 4.2 Demography

Once the geographical hierarchy has been built, we construct the virtual population. We initialize the population with age and sex attributes using statistical data from census records collected by UNHCR and the Government of Bangladesh for the camp Admin level 2 blocks (super areas) [28]. The number of residents in each IOM defined sub-block (area) is assigned proportionally according to population data collected at this level [42]. We naturally capture the heterogeneity in population density and demographic attributes by ensuring that our digital twin reflects the distribution of residents at the sub-block (area) level, and the statistical age and sex characteristics of the camp Admin level 2 blocks (super areas).

The final demographic attributes to be assigned to the virtual population are the comorbidities. An individual’s response to COVID-19 and other diseases can depend on the presence of comorbidities such as diabetes, heart conditions, and conditions causing immune suppression [43, 44, 45]. Since residents of the Cox’s Bazar settlement generally originate from Myanmar, we use national-level comorbidity distributions from Myanmar [46, 43, 47, 48], to probabilistically assign comorbidities based on the age and sex distribution of individuals in the settlement.^3^ In the specific case of COVID-19, given a lack of data and research on its impact on individuals with multiple comorbidities, it is assumed that an individual has at most one comorbidity. Our model can be adapted, however, once a greater body of literature is available. More details on the effects of comorbidities and their distribution can be found in Section 5.4 and Appendix E.

### 4.3 Shelters

Intra-family interactions create key transmission routes for infectious diseases. Correctly modeling family and shelter compositions is therefore important to enable realistic reproductions of infection modes. Data availability on household^4^ composition varies depending on the type of surveys and census carried out. Furthermore, in many settlements, multiple households may share a common shelter with little physical division between them. In settings such as the UK, detailed household composition data allows for a highly granular grouping of individuals into households [10]. In this case, we tailor our method of household and shelter distribution to the data available.

For the Cox’s Bazar settlement, we use data on the number of households at the sub-block (area) level as given by IOM [42], and data on the total number of residents in each Admin level 2 block (super-area) [28]. To account for data mismatches between the two datasets, we rescale the IOM’s household statistics so that the total number of residents estimated based on these household counts matches the total number of residents recorded in UNHCR’s census, at the Admin level 2 block (super-area) level. Once we know the (rescaled) target number of households for a given sub-block (area), we construct them by sampling each household’s size from the global household size distribution reported by UNHCR [49]. Each digital household is then populated according to the following algorithm:

1. We first allocate one adult (a person older than 16 years old) to each household, if available.
2. We iterate over all the non-full households, allocating one child (a person younger than 16 years old) each time until all children belong to a household.
3. For each non-full household, we choose an adult from the unallocated adult population by picking a person of the closest age to the current household adult resident, and of the opposite sex, if available.
4. The remaining adults are randomly distributed into households that still have space.

This algorithm naturally captures both single-headed households and child-headed households, as well as multi-generational families all of which are important for both protection concerns and disease spread.

In the left panel of Figure 2 we show a comparison between UNHCR data and the household distribution for the Cox’s Bazar settlement derived from our model. The distributions do not match perfectly due to the introduction of uncertainty when rescaling the number of households. Nonetheless, the two distributions are in reasonable agreement, with UNHCR reporting an average household size of 4.6 and our model producing a mean size of 4.4 (see the left panel of Figure 2).

**Figure 2:**
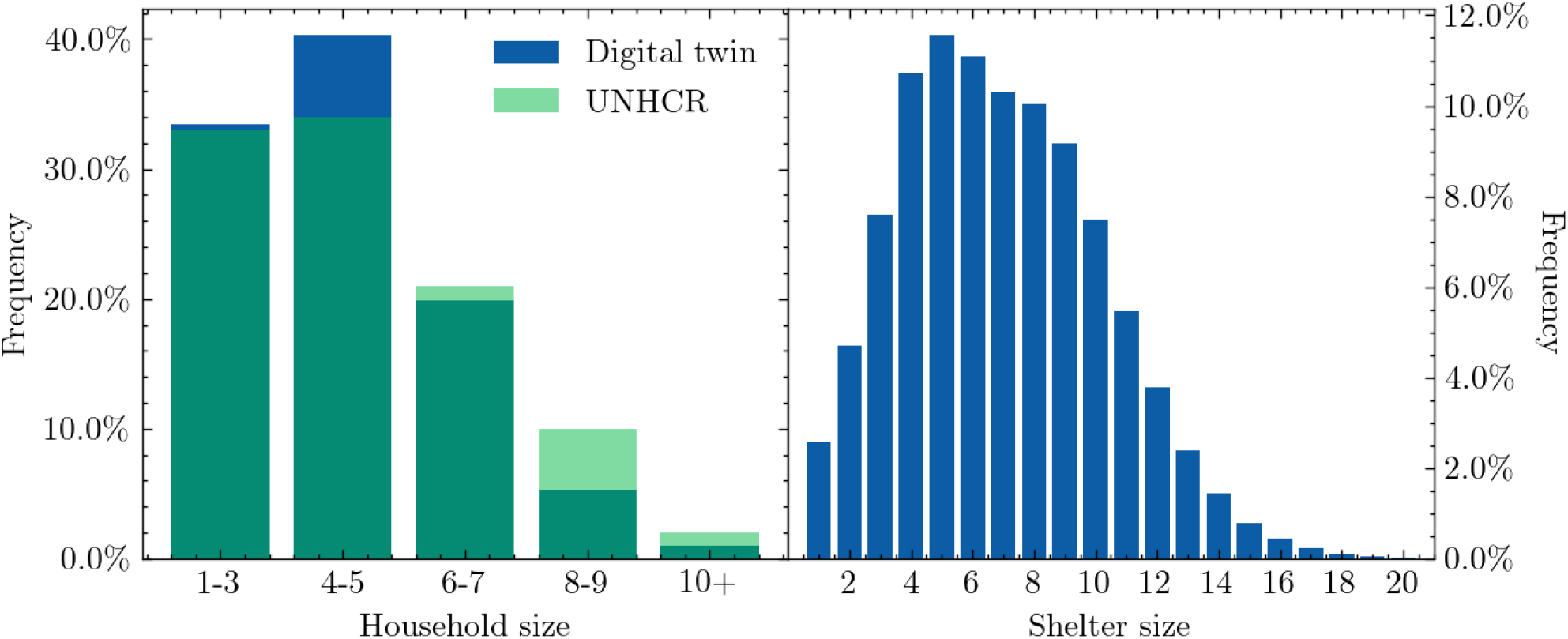
Left: Household size distribution comparing the UNHCR data with the digital twin of the Cox’s Bazar settlement, where the binning has been chosen to match UNHCR’s reporting structure. Right: Shelter size distribution from the digital twin.

Finally, once we have constructed the households, we group households into shelters. In the Cox’s Bazar settlement, approximately 75% of the families share a shelter with another family [5]. The resulting shelter size distribution is plotted in the right panel of Figure 2. The resulting average shelter size is 7 persons.

### 4.4 Learning centers

Learning centers refer to classroom settings for educating children and young adults [52]. Each settlement runs educational programs differently; therefore our base construction is designed to be flexible to account for such operational differences.

In the Cox’s Bazar settlement, due to the large number of children in the camps and the limited number of educational facilities, children usually attend the centers for two hours a day, and several blocks of two-hour teaching sessions occur daily in each learning center [54]. Children are assigned to a daily time slot in a specific learning center, during which - from a modelling perspective - they have the potential to interact with other children in the class, as well as their teacher. We assume that each learning center offers four two-hour teaching blocks per day. In our model, each classroom is assigned one teacher who is drawn randomly from the population in the area in which the learning center is located, and only children enrolled in the education system in the camp are sent to school (see Figure 3). Each enrolled child will attend the learning center closest to his/her shelter, unless that learning center has reached a maximum capacity of 35 pupils per shift, in which case the child will go to the next closest learning center. If the nearest 50 learning centers are all full, one of them is picked at random. The digital twin matches enrollment statistics collected at the camp (region) level, stratified by age and sex [50, 51, 52, 53]. Appendix B contains further details on how learning centers are constructed.

**Figure 3:**
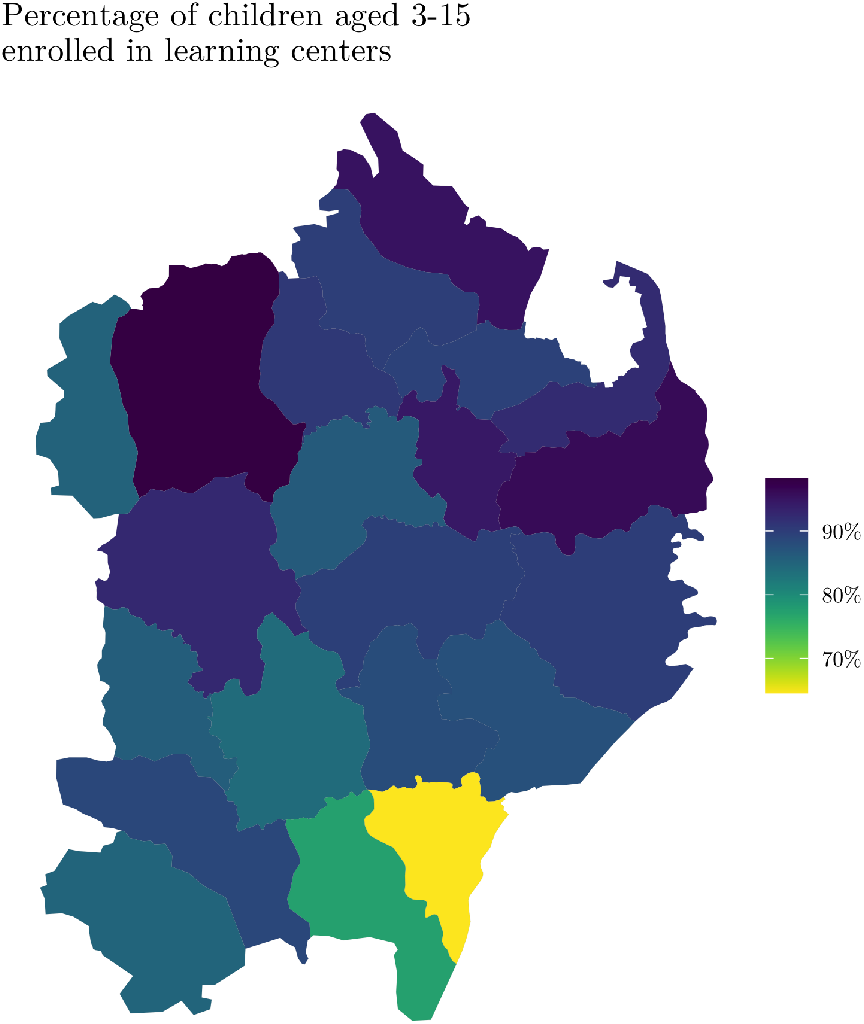
Enrollment of children aged 3-15 years old in each camp as a percentage of the total number of children living there [50, 51, 52, 53].

### 4.5 Dynamic Locations

The shelter and classroom constructions described above capture interactions between static groups of people. In other words, we currently assume that household and shelter compositions are fixed, along with the learning center attended by each enrolled child. However, there are many other locations at which attendance and mixing are highly dynamic, such as aid collection stations or visits to hand pumps and latrines.

Table 1 details the additional locations in the Cox’s Bazar settlement which we include in the model such as the markets where PoCs receive electronic vouchers which are used to purchase food products [55]. Using data collected by the Inter Sector Coordination Group (ISCG) [56], we place the dynamic locations using latitude/longitude coordinates as shown in Figure 4. Hand pumps and latrines are distributed to sub-blocks (areas) based on statistics detailing the number of individuals sharing such facilities [5], while play groups are dynamically created by randomly drawing together groups of children from neighboring shelters. We also model interactions between different shelters through family and individual visits. Each shelter is linked to up to 3 other shelters, and each member of a shelter can visit one of its linked shelters during a simulation time step. In Section 5 we discuss how we select which people visit these locations, and with what frequency, based on available research and literature. Appendix B contains further details on how these locations are constructed.

**Table 1:** Locations that simulated PoCs can visit in the model. We classify each location as being either indoor or outdoor.

| Activity | Type |
| --- | --- |
| Shelter Visits | Shelter |
| Distribution Center | Indoor |
| Non-Food Distribution Center | Indoor |
| E-Voucher Outlet | Indoor |
| Communal Center | Indoor |
| Safe Space for Women and Girls | Indoor |
| Religious Center | Indoor |
| Learning Center | Indoor |
| Hand Pump and Latrine | Outdoor |
| Play group | Outdoor |

**Figure 4:**
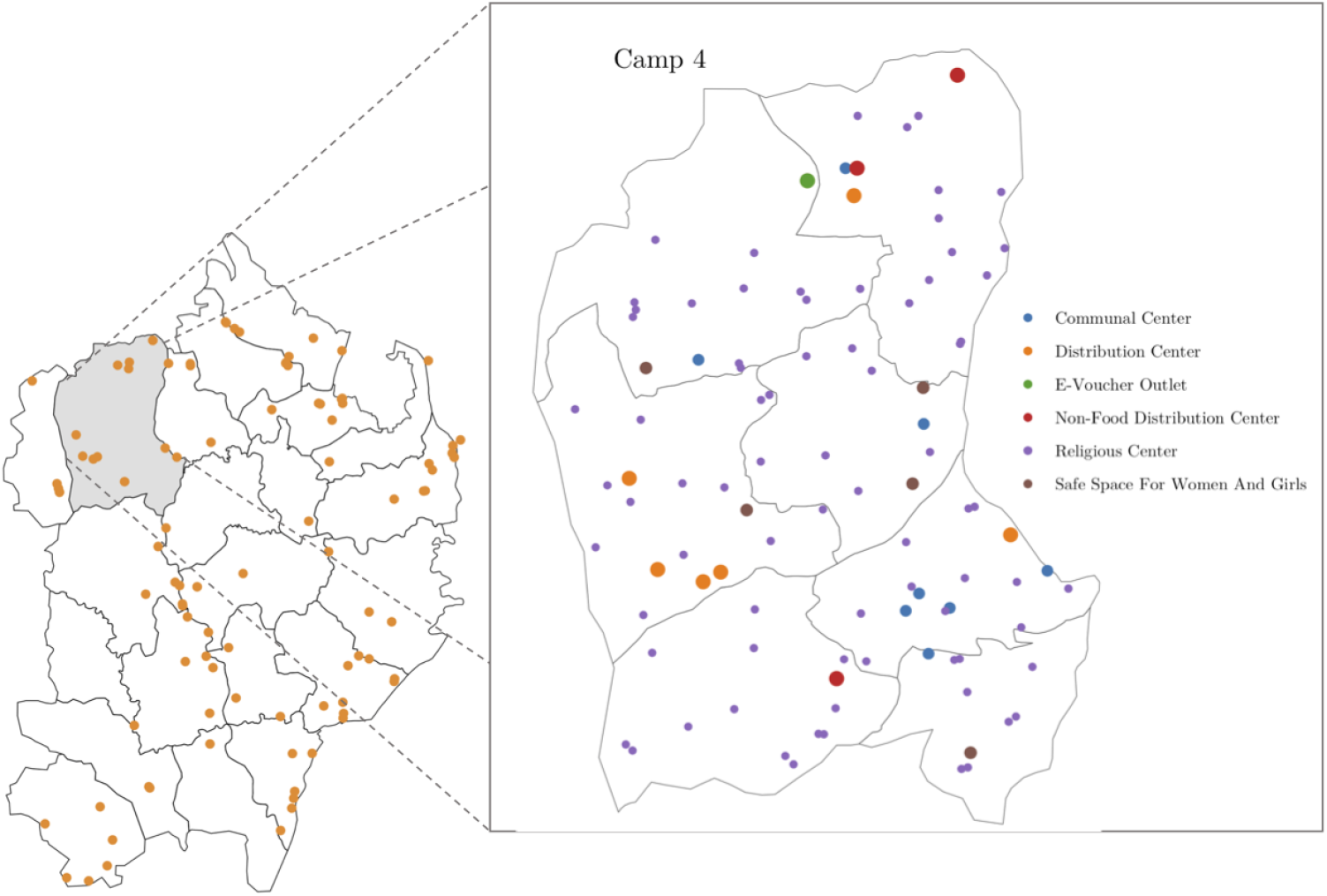
Left: locations of modeled distribution centers in the settlement. Right: Zoomed in view of Camp 4 showing six different kinds of modelled locations.

## 5 Simulator

The second stage of our modeling process involves designing the simulator which probabilistically models the social mixing and dynamic interactions of the virtual population. The digital twin forms the basis upon which the simulator is constructed. Each person in the model has the potential to move and interact with others based on individual and group dynamics which are derived from data. Since we model movement at the individual agent level, we have the ability to flexibly change all parameters used in the model and allow for different social mixing behaviors. In this section we describe the setup in general terms and its specific application to the Cox’s Bazar settlement.

### 5.1 Daily routine

To model the movement of individuals in the camp, we divide each simulated day into several time steps as shown in Figure 5. In each step, an individual has a certain probability of doing one of several possible activities, during which they might interact with others who carry out the same activity in the same location contemporaneously. If no activity is chosen the agent will remain in their shelter.

**Figure 5:**
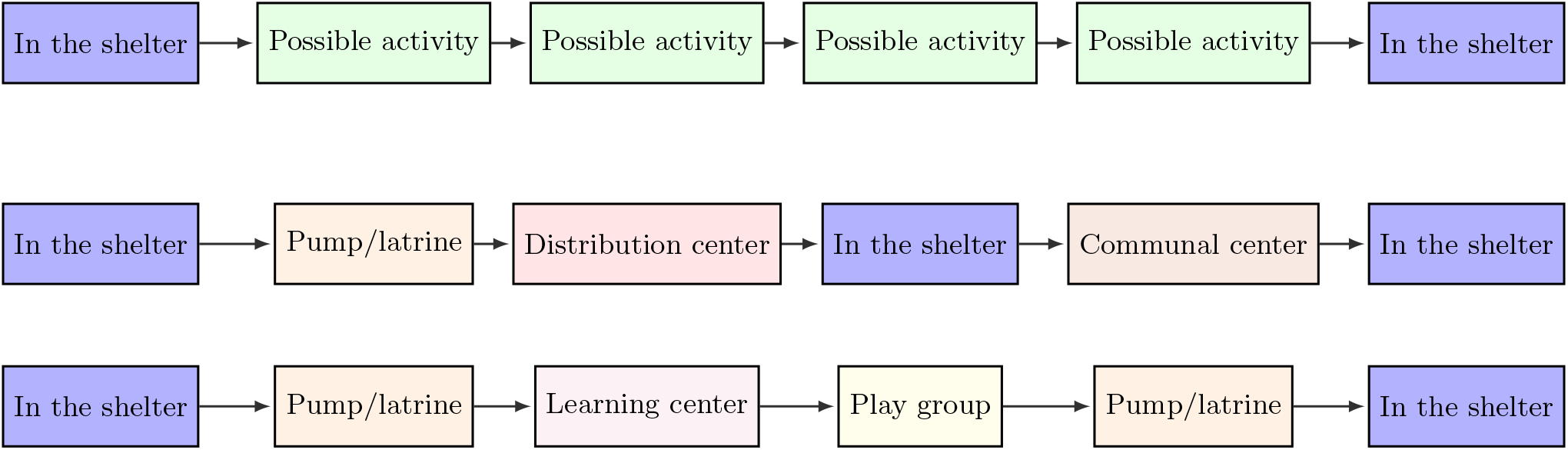
Top: Daily routine structure for individuals modeled in the simulation. We allow each individual to perform up to four possible activities per day (although this can be flexibly changed). If an activity is not chosen, the individual returns to their shelter. Middle: Example of a simulated day for an adult in the settlement. Bottom: Example of a simulated day for a child attending a learning center in the settlement.

Simulated individuals begin the day in their shelters, where they can interact with others in their household, or other households which share the same shelter (see Section 4.3). After this time step, each day contains four or five opportunities for the individuals to undertake an activity in one of the locations listed in Table 1. The probability that an individual carries out a certain activity is determined based on their age and sex and available research on individual activity patterns (see Appendix B for more details). This implementation enables realistic modeling of how individuals behave as we can tailor the amount of time individuals spend in a given location, and how these probabilities depend on demographic attributes of the population.^5^ When choosing which location of a given activity to attend, e.g. which communal center to attend, individuals in the model are given a choice of their nearest 5 locations corresponding to the relevant activity.^6^ This choice allows for the capturing of both local and inter-camp mixing.

To choose the activity each individual participates in at each time step, we follow the same methodology as used in JUNE [10]. Each individual is assigned a Poisson parameter according to their age and sex, *λ*, and the probability that an individual does a certain activity during a given time step is determined through a Poisson process. ^7^

To assign activities to individuals, we first check if the individual does any activity at the given time step:

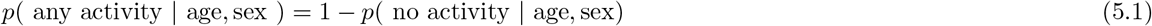

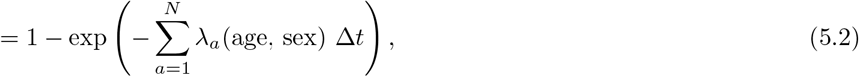

where *λ*_*a*_(age,sex) is the Poisson parameter associated with activity *a* for a person with a given age and sex, *N* is the number of possible activities and Δ*t* is the amount of time allowed to do a given activity (currently this parameter is fixed at 2 hours, however can be varied if necessary). If no activity is performed then the individual returns to their shelter.

If a person carries out an activity, the next step is to determine which specific activity is chosen. The probability that activity *a* is chosen given that the person does any activity is given by:

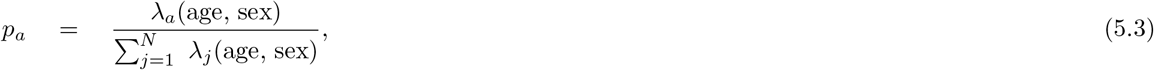

Once an activity has been determined for a given individual, they are moved to the relevant location where they can interact with others who are also present.

### 5.2 Transmission

At each time step, different collections of individuals will inhabit the same space (e.g., a distribution center or play group), which we will refer to as a ‘group’. Each group is a collection of individuals in the same geographical location performing the same activity. Individuals in the same group have the opportunity to interact with others in the group. If one or more of the people in the group are infected, there is a chance that a susceptible individual may become infected. We implement the transmission dynamics described in Bullock et al. [10], which we briefly outline here for completeness.

For a susceptible individual, *s*, we define the following properties: *L* is the location type (e.g., distribution center); *g* is the set of people present in a specific location; *i ∈ g* is the subset of infectious people in that location; *t* is the current time step; Δ*t* is the length of time that individuals remain in the location; *Ψ*_*s*_ is the susceptibility of individual *s*; *ℐ*_*i*_(*·*) is the infectiousness of individual *i* over the time frame *t* to *t* + Δ*t*; 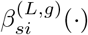 characterizes the intensity of contacts between individuals *s* and *i* at a point in time; and *𝒫*_*s*_(*·*) is the probability that susceptible individual *s* is infected over a given period of time.

The probability of a person, *s*, being infected in the time step [*t, t* + Δ*t*] is taken to be:

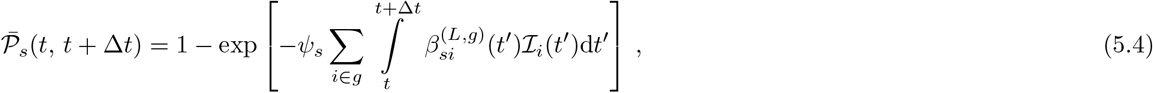

As expected, staying longer in a location, being more susceptible to the disease, being surrounded by more infectious people, and having more intense contacts all increase the probability of disease transmission.

The intensity of the contacts, *β*, depends on the location, the number of people in the group, time, and the number of contacts along with the proportion of which are physical (as opposed to less intense conversational contacts). To represent this, the intensity of contacts between susceptible individual *s* and infectious individual *i* who are part of group *g* in location *L* at time *t’* is taken to be:

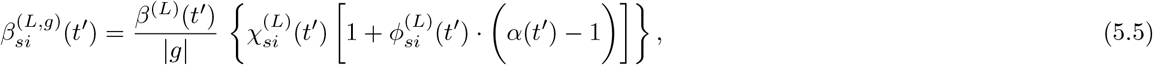

where |*g*| is the number of people in group 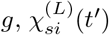 is the number of contacts between people with the ages of individuals *s* and *i* over a given time period, 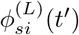 is the proportion of those contacts that are physical, and *α*(*t*^*’*^) describes the additional infection risk of the physical contacts relative to the conversational ones.

In an ideal situation, the mixing matrices 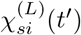 would be derived from survey data [58, 59, 60], but such data has not been collected in the settlement setting.^8^ Instead we use a mixture of survey and observational data, along with various assumptions to hypothesize mixing matrices such that they capture the broad structure of contacts in a given location while allowing the values of the *β*^(*L*)^(*t’*)parameters to absorb the details of the precise number of contacts. A similar argument can be made for the values of 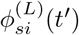 Since *α*(*t’*) is fixed for all locations, this just serves as a scaling parameter for the physical interaction matrix and so can be arbitrarily set. We heuristically take *α*(*t’*) = 4 to account for increased intensity likely found in the settlement environment.^9^ We detail our choices for 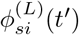 and relevant assumptions in Appendix C.

For modeling the spread of COVID-19, we use the time-dependent infectiousness profile, *I*_*i*_(*t’*), presented in [57] and represented in Figure 6. As in [10], we reduce the infectiousness of asymptomatic individuals, relative to that of a symptomatic individual, by a factor of 0.5.

**Figure 6:**
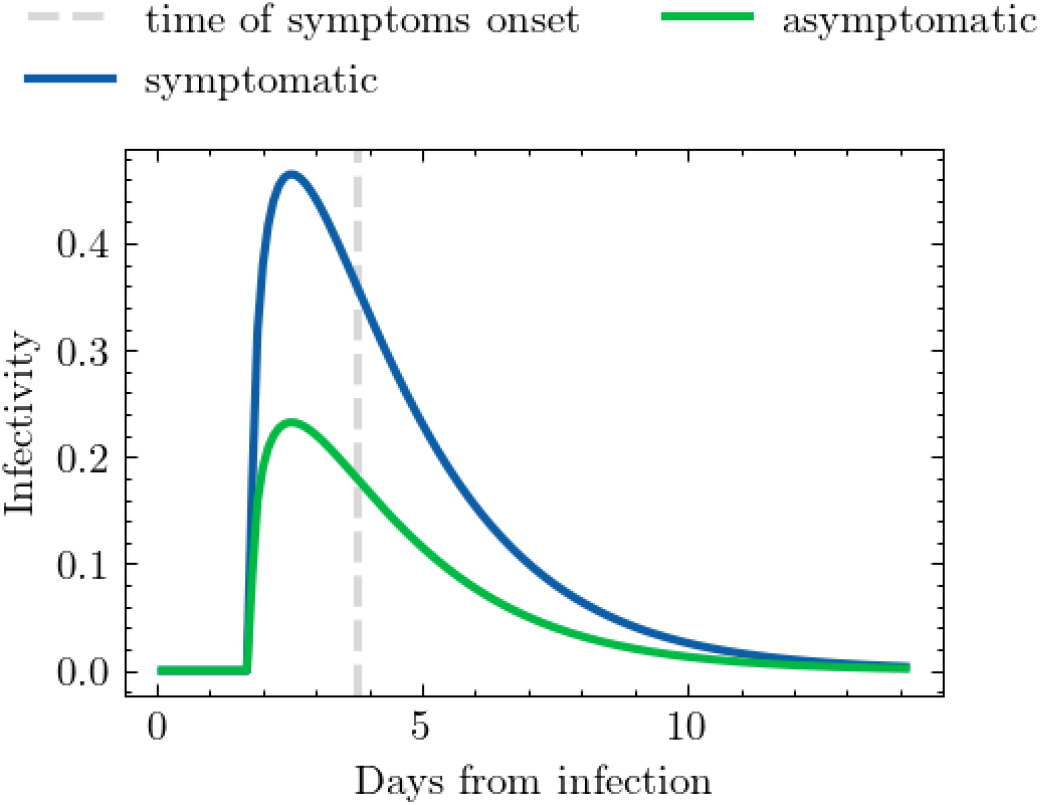
Infectiousness profile as a function of time derived from [57].

### 5.3 Disease progression

Once an individual has been infected, we assign them a health trajectory that controls the severity of their symptoms as a function of time. Each trajectory is determined by the final outcome of the disease (whether the individual recovers or dies), the different stages over which the individual will go to arrive at that final outcome, and the duration of each stage. In Figure 7, we show the stages that we include, together with the possible trajectories defined by arrows. We incorporate the distinction between mild and severe individuals to differentiate between people that show symptoms but are still well enough to leave their home, and those that developed a more severe illness that prevents them from doing so. An important factor hard to account for when estimating the probability of trajectories that involved hospitalization is the evolution and diversity of healthcare services seeking behaviour [12], however we leave this as a line of future work due to the lack of publicly available data for the specific case of COVID-19.^10^

**Figure 7:**
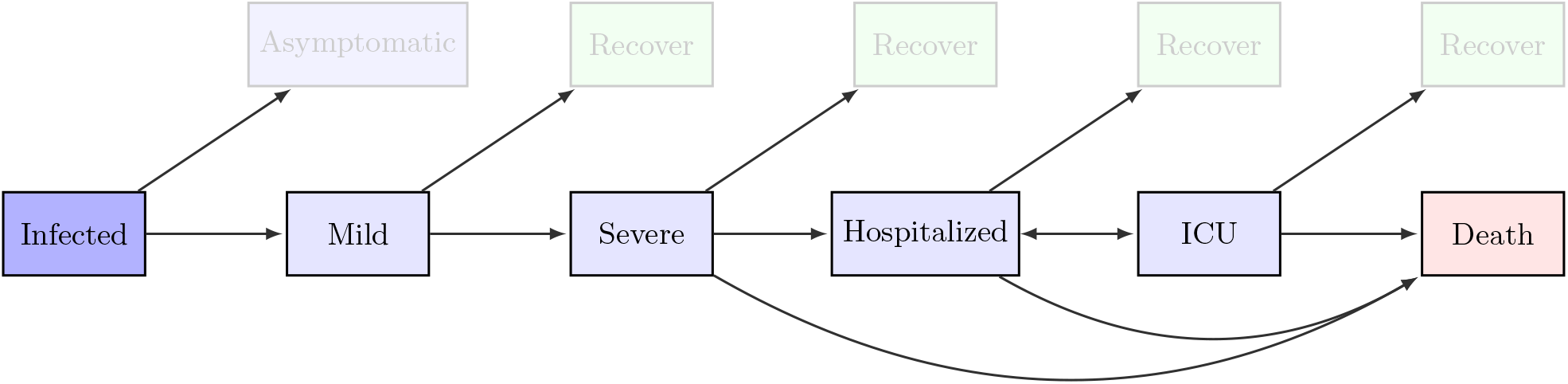
Possible health trajectories modeled. When an individual is infected, they might remain asymptomatic, or become symptomatic with symptoms likely to progress. In our implementation, the likelihood of each of these transitions is dependent on the age, sex, and/or comorbidities of the infected individual. As more data becomes available, additional factors can also be included.

The probability to follow one of the outlined trajectories depends primarily on the age and sex of the infected individual. We use estimates from [10], derived from UK data, to determine the likelihood of each trajectory for the particular case of COVID-19. However, translating hospitalization and mortality rates from one country to another is not straightforward. Firstly, it is not clear to what extent these estimates are influenced by the capacity and quality of the healthcare system from where data is taken from. Secondly, the prevalence of chronic illnesses can increase the risk of severe illness in response to COVID-19 [43]. Whereas the first aspect requires further research to correct for, we have included a correction to the effect of comorbidities prevalence in the Cox’s Bazar settlement, following [48], that will be explained in detail in the next section.

### 5.4 The effect of comorbidities on disease progression

To better reflect the specific evolution of the virus in the Cox’s Bazar settlement, we accounted for location-specific age, sex, and comorbidity distributions which are assumed to affect the probability of severe infection. As the PoCs in the settlement are originally from Myanmar, national distributions of comorbidities by age and sex from Myanmar were used along with demographic data on the distribution of age and sex from the settlement itself (although the limitation of this use of national level data in the context of marginalized populations was noted in Section 4.2). Comorbidities were assigned to the virtual population based on this data; with the assumption, for practical reasons, that an individual has at most one comorbidity that would affect the probability of a severe infection. The probability of severe infection adjusted for comorbidity, age, and sex was then used to determine the individual’s disease progression if infected (further details are found in Appendix E).

In Figure 8, we show the increased risk of severe infection after accounting for comorbidities relative to the health trajectory probabilities modelled for the UK [46, 43, 47, 48]. We define the increase risk as the mean probability of a severe outcome (severe symptoms, hospitalization, or death) for a population with the comorbidty prevalence in the Cox’s Bazar settlement, divided by the mean probability of a severe outcome for a population with the comorbidity prevalence of the UK. The effect is most important for ages between 20-70.

**Figure 8:**
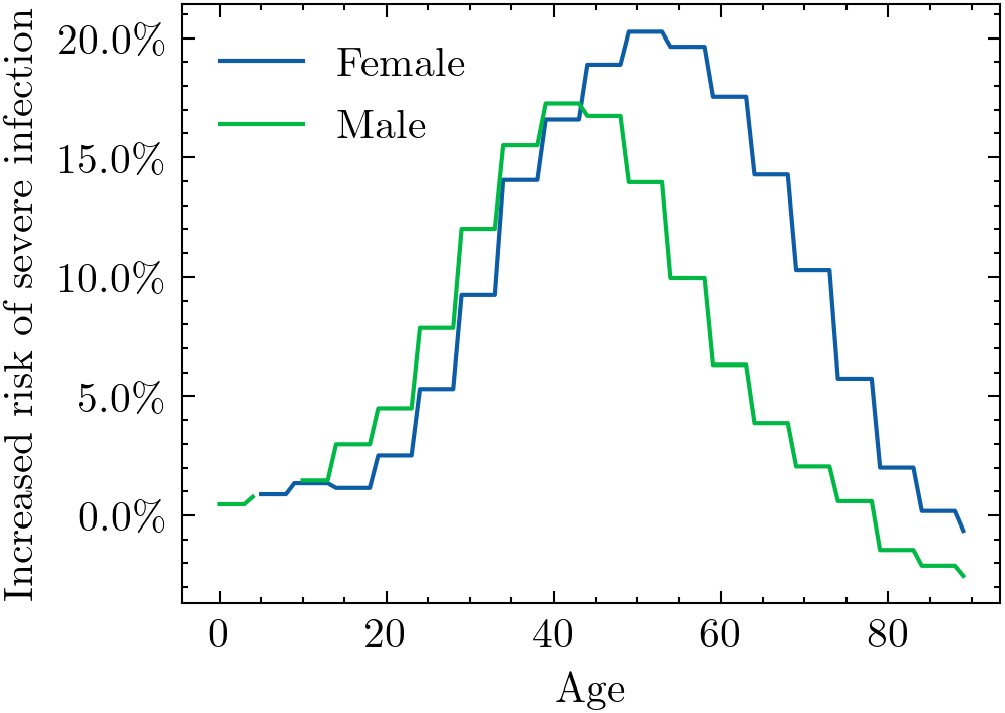
Severe infection rates adjusted for estimated comorbidities in the PoC population using as base-line UK data [46, 43, 47, 48].

## 6 Modelling Operational Interventions

Modelling the effectiveness of different operational interventions such as alternative home care delivery mechanisms, mask wearing, and (re)opening learning centers is important for future planning purposes. In a refugee settlement, implementing such interventions requires significant advance mobilization as well as operational and financial support. This is especially true in the Cox’s Bazar refugee operation, where the language of the primary beneficiaries (the Rohingya) does not have a written script, meaning that communication can be challenging [61]. By simulating the possible effects of operational interventions prior to their introduction, and incorporating the results of these simulations into decision making processes, intervention priorities can be better identified.

Once the digital twin and simulator have been set up, we are able to run simulations under different parameter configurations. As mentioned in Section 1, in a context of incomplete testing and case reporting data, we focus primarily on analyzing intervention efficacy through comparing the relative magnitude between different implementation conditions. Different models and approaches can account for different degrees and types of uncertainty making consensus on statistical predictions challenging even in more data rich environments. However, despite often highly variable predictions, consensus can often be reached on ranking intervention efficacy [62] which can be of interest for decision making.

Since we take an agent-based approach, operational interventions are typically implemented in one of two ways. First, they can be implemented as changes in simulator parameters such as the intensity of interactions in different locations, which can correspond to interventions such as mask wearing. Second, we can dynamically adjust the movement of individuals in the model, which can correspond to the closure of certain amenities and/or changes in their functionality.

Our model allows for flexible experimentation with different operational interventions. Below, we summarize results from several simulations which were of particular interest in the COVID-19 decision-making context. In particular, we compare:

1. The impact of encouraging patients with mild symptoms to immediately self-isolate at home (“home-based care”), relative to presenting themselves at isolation treatment centers after various time delays from symptom onset.
2. The impact of variations in the efficacy of, and compliance with, mask wearing in difference locations.
3. The impact of opening child learning centers and possible ways to mitigate its potentially negative consequences on the epidemiological development of the disease.

The primary metrics we use to assess and compare scenarios include the time to infection peak, the height of the peak, and the total number of infected. Time to infection peak and the height of the peak are important because they serve as a proxy for how quickly a settlement’s response capacities will be overwhelmed and to what degree; all else equal, responders would prefer a slower rise in infections in order to have more time to prepare for a surge. Total infections are important because they are a proxy of the settlement-wide impact of COVID-19.

### 6.1 Epidemic spread baseline

Before we can assess the effects of interventions, we must have a baseline model of disease spread against which to compare. In data rich environments such as the UK [10], epidemic models can be fitted with high resolution data on confirmed cases, hospital admissions and fatality statistics. However, in the Cox’s Bazar settlement, due to limited COVID-related statistics, fitting the model to data with a high degree of confidence is not possible. Initially this was due to a lack of testing facilities, however, as more testing has become available, other factors such as rumors and misinformation have driven under-reporting and discouraged potentially infected individuals from coming forward [63].

As mentioned in Sections 4-5, our model of the Cox’s Bazar settlement is largely constructed using input data and parameters derived from surveys and other reports regarding its population and the epidemic. We use the intensity parameters (i.e. *β*^(*L*)^ parameters - see Equation 5.5) to adjust the strength of interactions, and therefore the probabilities of transmission, in different locations. To construct a baseline simulation against which to test operational interventions, we focus on estimating the relative magnitudes of these location dependent parameters based on a mixture of the limited current case reporting data and heuristics from the literature.

Intensity parameters are categorised into three groups according to the locations they correspond to: shelters, indoor and outdoor settings - as defined in Table 1. Transmission in the home environment is one of the lead drivers of disease transmission and therefore we first estimate the intensity parameter in this setting. Details on how we perform this estimation can be found in Appendix D.2. Depending on the type of intervention being tested, the indoor and outdoor intensity parameters are varied relative to the estimated shelter parameters according to their importance to that specific intervention. In certain scenarios, to limit the number of concurrent parameter variations, we fix the ratio of the intensity parameters in indoor settings to that of the shelter to 55%, and the ratio of intensity parameters in outdoor settings to that of the shelter to 5% as discussed in Appendix D.2. This heuristically captures the relative differences in the types of contacts made in these locations, along with the reduction in transmission probability in outdoor environments suggested in the literature [64, 65, 66].

When developing the baseline infection model, we aim to include the relevant policy decisions currently implemented in the settlement. For example, since March 2020 the Safe Spaces for Women and Girls (SSWGs), and learning centers have been closed in an attempt to mitigate intermixing and viral transmission [67, 68]. These closures are incorporated into out model. Similarly, aid collection from distribution centers can occur up to twice per week for each household, however, the World Food Programme (WFP) has implemented policies to reduce this to once per month and converted several non-food distribution centers to serve as distribution centers as well [55, 69, 70]. In addition, E-voucher outlets, which would normally operate as market places where PoCs receive vouchers to purchase food, have been converted to operate as distribution centers [69, 70]. We therefore alter the probabilities that individuals go to distribution centers and E-voucher outlets in line with these regulations. Appendix B details further the assumptions made for each activity that a digital person may participate in.

For simplicity, in the baseline model we assume that all symptomatic individuals with mild symptoms self-quarantine in their shelter with a low compliance of 30% to account for difficulty in communicating quarantine procedures, as well as the inability of many individuals to properly quarantine given certain basic needs [71]. As mentioned in Section 5.3, those with severe symptoms are by definition required to stay in their shelter in line with [10]. Given the makeup of the shelters, quarantining individuals will not leave their shelter but may still interact with those in their shelter. Those with severe symptoms are always assumed to stay in their shelter by definition (see Section 5.3).

All models are seeded with 88 infected individuals across a variety of geographic regions based on data collected on the 24th May 2020 [13]. Given this choice of seeding by location, our baseline model was found to reproduce the geographic spread of the virus in its early stages within reasonable agreement. However, use of data for comparison beyond this initial phase is challenging due to the limitations mentioned previously. Details of the rationale and precise procedure for the initial seeding is given in Appendix D.1.

### 6.2 Isolation centers

In many countries, those with symptoms which are not yet severe enough to require hospitalization are encouraged to stay at home and self-quarantine. In the case of settlements such as that in Cox’s Bazar, the density and living conditions of the residents mean that avoiding contact with family in the home environment is not possible, and individuals frequently have to leave their shelter to use facilities such as hand pumps and latrines. In an attempt to better enable the isolation of symptomatic individuals, public health officials in the settlement set up isolation and treatment facilities to house those who have tested positive for COVID-19 but do not require hospitalization [72, 73].

Given the limited capacity within the settlement’s isolation and treatment centers, and given the likelihood of a high-transmission scenario in the settlement, isolating every symptomatic PoC may rapidly exceed the capacity of these centers. Thus, we modelled an alternate scenario of home-based care for PoCs with mild and severe symptoms thereby allowing these centers to provide additional capacity to the regional hospitals. Specifically, we modeled a scenario (a) in which patients with mild and severe symptoms (not requiring hospitalization) self-quarantine and are treated at home (we refer to this scenario as “home-based care”). A schematic of this scenario is given in Figure 9. In contrast, we also modeled a scenario (b) in which symptomatic patients go to isolation and treatment centers regardless of symptom severity up until they need to be hospitalized (we refer to this scenario as “treatment center scenario”).

**Figure 9:**
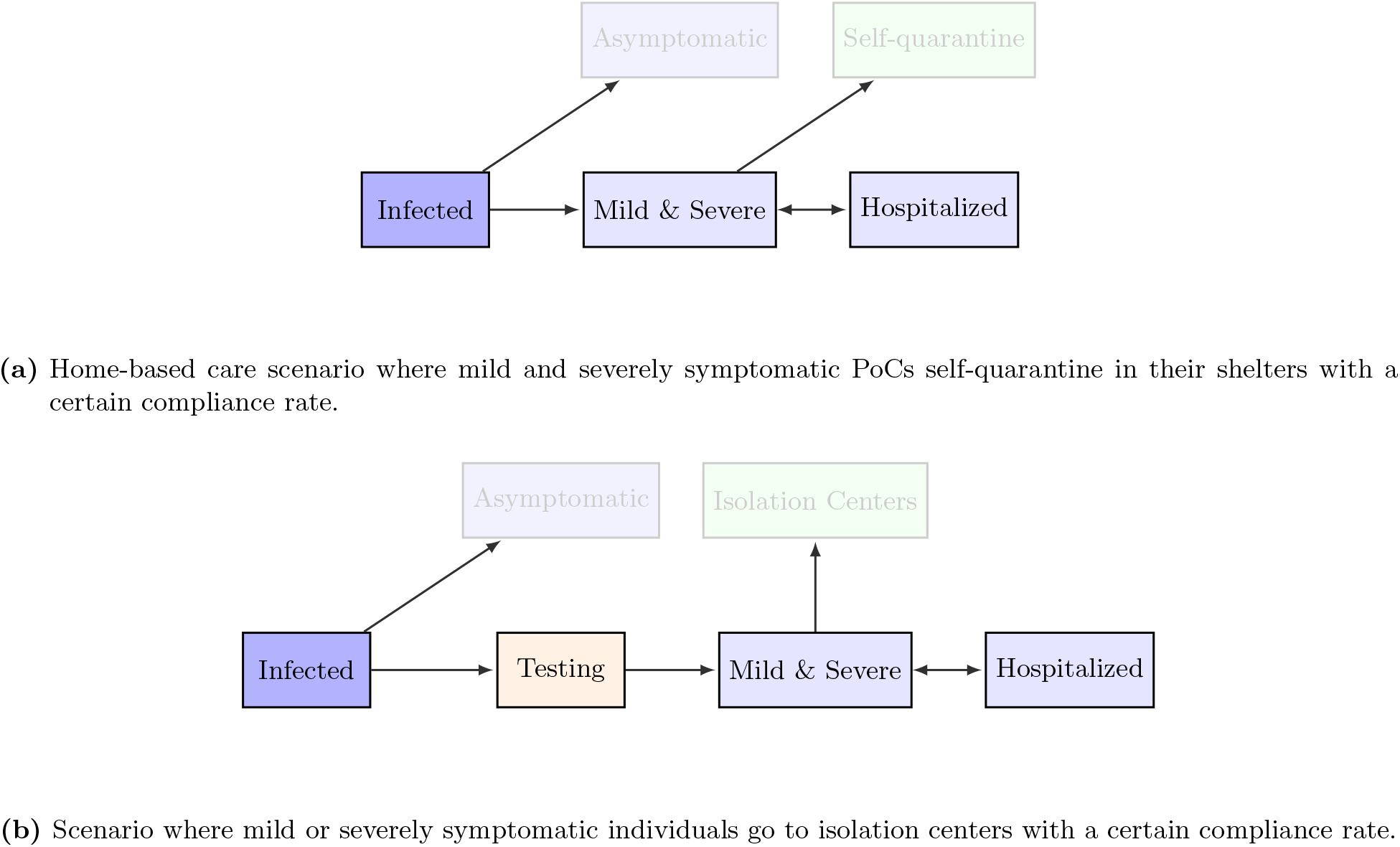
Possible simulated care trajectories modelled.

To explore different treatment center scenarios, we varied the average time delay between symptom onset and isolation - this is designed to encapsulate the delay between a symptomatic individual developing symptoms and presenting themselves for testing, as well as the time taken to process the testing - as well as the the time spent in the isolation center. The former was varied between 0-5 days while the latter was varied between 5-10 days. Further details on these parameters are given in Appendix F.1.

Figure 10 presents the outputs of our simulations under the different scenarios tested where the baseline denotes the home-based care scenario. Since the key concern in introducing a home-based care plan is the potential increase in the number of cases given the continued interaction with those in the shelter environment, we show focus on the rate of daily infection as well as the percentage of the population infected.

**Figure 10:**
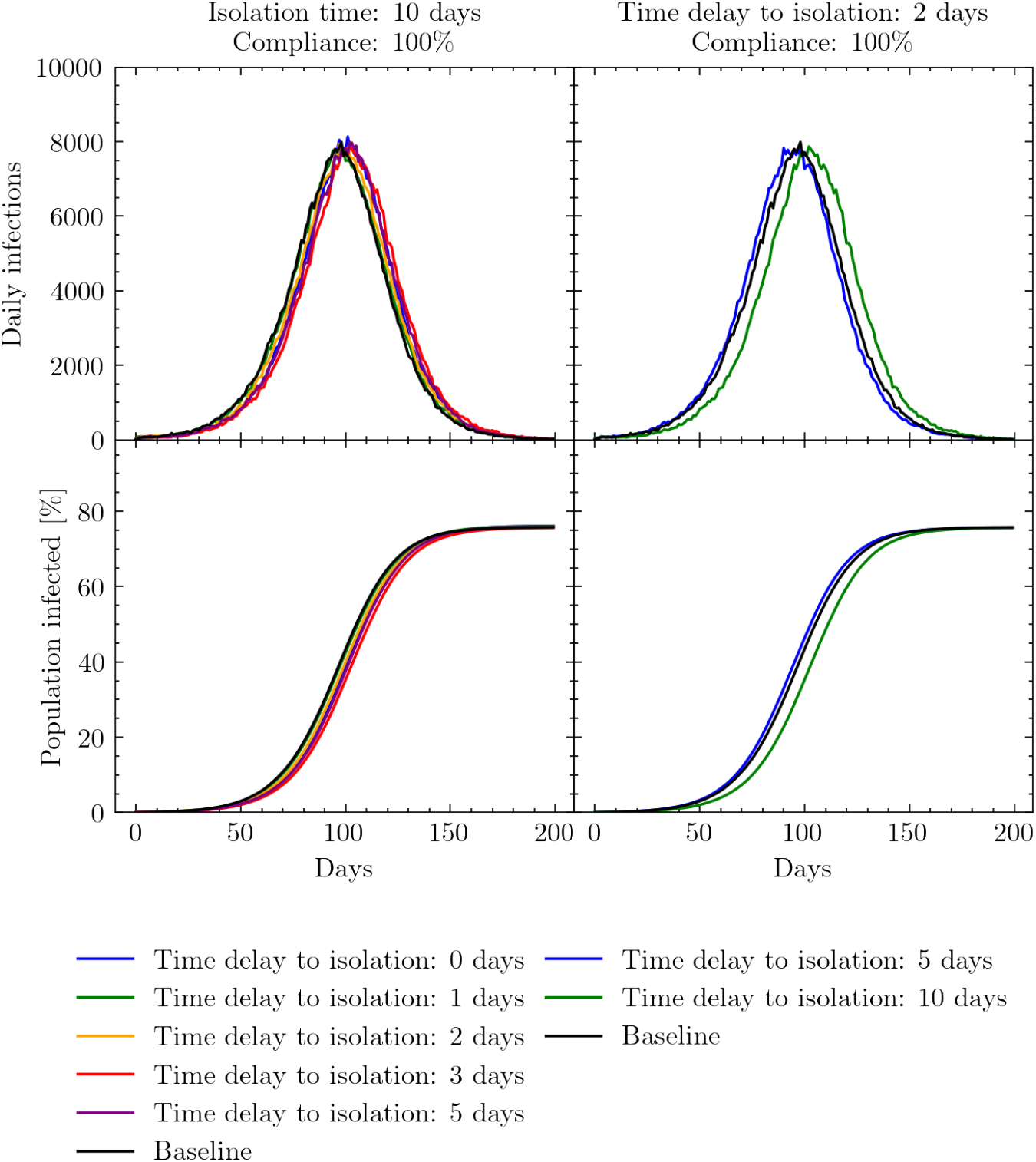
Simulated daily and cumulative infections measured in days since the beginning of the simulation. Left: exploring the effects of varying the mean time delay to isolation from symptom onset relative to the baseline home-care scenario where mild and severe cases are treated at home. Here, the time spent in the isolation and treatment center is fixed at 10 days. Right: exploring the effects of varying the time spent in isolation while keeping the mean time delay to isolation fixed at 2 days. In both scenarios the compliance rate that people present themselves for isolation is set too 100%.

We first simulate, the effects of varying the average time delay between an individual developing symptoms and presenting themselves at an isolation and treatment center. To disentangle parameter dependencies, we fixed the time spent in isolation to 10 days and the compliance level for an individual to go to the centers at 100%. This presents a best case scenario for the isolation of individuals. From Figure 10 we see that varying the average time delay to isolation had little effect relative to the baseline home-based care scenario.

Secondly we simulate the effects for varying the time spent in the isolation center, while fixing the compliance to 100%. Here we also fix the average time delay to isolation to 2 days to represent an optimistic yet realistic scenario. Again, we see this has little effect on the daily infection rate relative to the home-based care scenario.

The reason for this observed lack of difference between the treatment and home-case scenarios is likely due to the infectiousness profile presented in Figure 6. Given the likely rapid transmission of infection between those residing in the same shelter, the majority of infections have likely taken place before symptom onset. To test our sensitivity to this profile, we examined the effects of shifting the peak of the infectiousness profile *±* 10 % but this yielded little difference in results. We do not present results at lower rates of compliance as these ‘best-case’ scenarios demonstrate relatively little effect.

The results presented in this section suggest that encouraging home-based care for individuals with mild symptoms may not have a major negative impact on the number of daily infections as was the initial concern. Therefore, if needed, it may be worthwhile to use isolation and treatment centers to supplement hospital bed capacity while encouraging those with less severe symptoms to isolate as best as they can in their shelters.

### 6.3 Mask wearing

Widespread adoption of face masks has the potential to significantly reduce the transmission of COVID-19 [74, 75]. In the settlement, surgical and cloth masks have been distributed to many PoCs, with the majority being of the latter type [70, 76]. The overall success of mask policies are contingent on both the efficacy of the masks themselves and compliance with mask wearing. To test the potential effect of mask wearing on the epidemiological development of COVID-19, we simulate the wearing of masks of different efficacies in all settings outside the shelter, with the exception of play groups, with variable rates of compliance. As in [10], this is encoded through a change in the interaction intensity parameters which are adjusted according to:

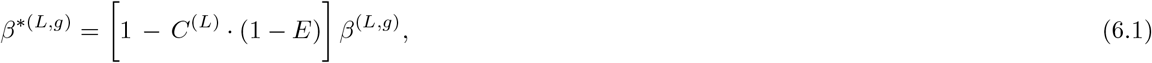

where *β*^*\**(*L,g*)^ is the new interaction intensity parameter, *C*^(*L*)^ is the compliance with correct mask wearing in a given location *L*, and *E* denotes the mask efficacy. Efficacy is defined as a function of the mask material, as well as any degradation through incorrect reuse and washing.

For the simulations described below, mask efficacy and mask wearing compliance were varied. For simplicity, the total compliance was varied equally across all locations, however compliance rates stratified by location can be tested in the future if necessary. More details on mask wearing efficacies and our parameter choices can be found in Appendix F.2.

Figure 11 shows the simulated effect of mask wearing on the daily number of infections as a function of compliance and mask efficacy. When mask efficacy is low, e.g. of the order of 20%, relative changes in compliance have a comparably small effect on the total proportion of the population infected. As the average efficacy of the masks increases, these changes in compliance can have a clearer effect, yet we see that further increases in average mask efficacy beyond the 50% level may have diminishing returns in realistic scenarios. For example, obtaining masks with average efficacies greater than 50% may be challenging and costly, especially when efficacy is also a function of correct mask useage, and the resources required to achieve this may be greater than the gain in transmission reduction.

**Figure 11:**
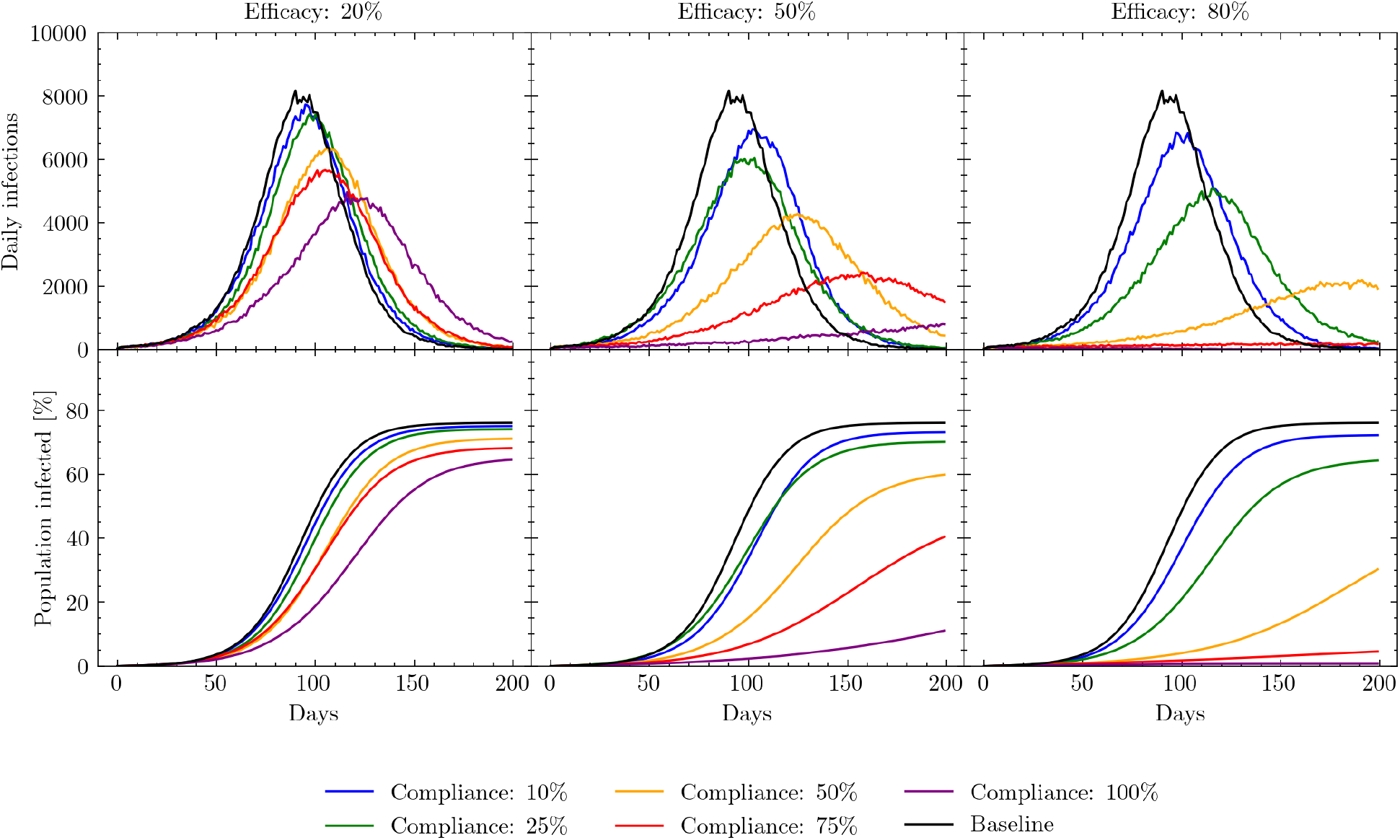
Simulated daily and cumulative infections measured in days since the beginning of the simulation. Results show the effects of varying the compliance with mask wearing in different locations under different assumptions regarding mask efficacy. The baseline model is the scenario in which no masks are worn.

As discussed at the beginning of this section, assessments of both the peak location and the total number of individuals infected can provide useful operational insights. Figure 12 illustrates how varying both compliance and efficacy affects both daily infection peak height and the total proportion of the population infected. As expected, both the height of the daily infection peak and the total number of people infected decline as mask efficacy and compliance increase. However, this figure also shows that peak height and total number of infections respond differently to changes in compliance or efficacy: peak height reduces faster than the total number infected as compliance and efficacy increase (i.e, it is more sensitive to changes in compliance and efficacy).

**Figure 12:**
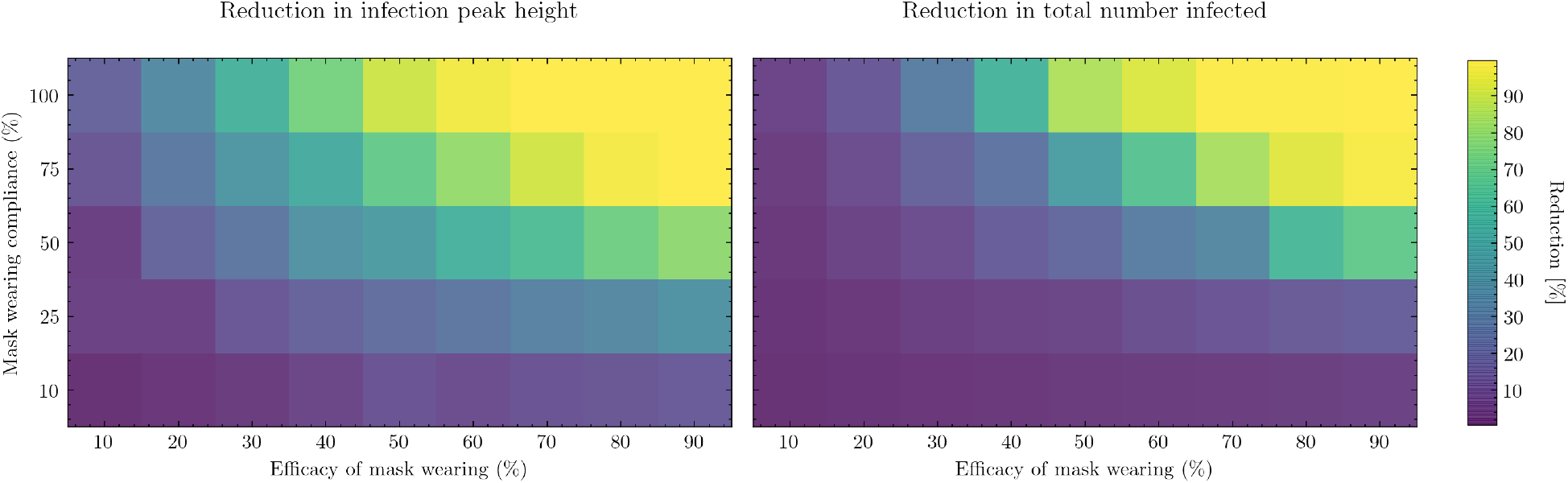
Left: percentage change in daily infection peak height as a function of mask wearing efficacy and compliance relative to the peak height of the baseline model. Right: percentage change in total number of infections up to a fixed point in simulated time as a function of mask wearing efficacy and compliance relative to the total number of infections simulated by the baseline model. The baseline model assumes no masks are worn.

In the case of mask wearing specifically, it is informative to know where to utilize already scarce resources - on increasing compliance or on increasing the average efficacy of the mask. From Figure 12 we also see the relative sensitivity to the compliance and efficacy parameter individually. For example, assuming an attainable efficacy of 50% [77] with a compliance rate of 50%, it can be seen that an increase in compliance by 25% has a similar effect to increasing mask efficacy by 30% on the daily infection peak height. Such considerations are operationally important as the wider use of lower efficacy masks which can be homemade and reused, thereby increasing compliance, may be considerably easier than importing large quantities of higher quality single use masks, such as surgical masks. See Appendix F.2 for more details on mask wearing efficacies and parameter choices.

### 6.4 Opening learning centers

Learning centers in the settlement have been closed since March 2020 in an attempt to mitigate the spread of COVID-19 [68]. However, keeping them closed can have serious consequences on the educational development of the children as well as increase concerns around child protection. Although opening learning centers imposes a risk of infection within the classroom, the closure of learning centers may also have negative consequences on the epidemiological development of the virus: when children are not in school, they participate in various activities such as assisting with aid collection, going to communal centers, or meeting up with other children and playing in groups outside which all serve as additional channels for intermixing [51, 52, 53]. Indeed, since the learning centers have been closed, settlement officials have observed an increase in children meeting up and playing in small groups [71].

To simulate the possible effects of opening the learning centers, relative to a set of baseline models in which they remain closed, we allow all children enrolled in the education system to go to school each day as described in Section 4.4. To avoid multiple concurrent parameter variations, in previous intervention scenarios we held fixed the interaction intensity parameters in different locations as the relative sizes of these parameters were not as important as others to understanding the potential effects of the intervention. However, in the case of learning center opening the relative intensity of interactions in the indoor and outdoor environments becomes key since when children are not in learning centers they are predominantly in outdoor environments.

Currently, it is unclear how intense interactions in learning environments might be relative to interactions with other children outside. To account for this unknown relationship, we varied the ratio of the interaction intensity in both indoor and outdoor settings to the interaction intensity in shelter settings while preserving the shelter-indoor-outdoor intensity hierarchy described earlier in this section.

Figure 13 shows the effect of opening learning centers on the daily and cumulative number of infections. The left set of panels demonstrate that varying the indoor intensities can have a non-trivial effect on the progression of the virus through the population although the two scenarios - opening the learning centers or keeping them closed - remain well distinguished from each other in both peak height and timings. However, the right set of panels show that varying the outdoor intensity can have significant effects on both peak height and location, with some scenarios less well distinguished. This difference occurs as, with the exception of learning centers, indoor locations outside of the shelter environment are much more irregularly visited by children in comparison to the rate at which they meet up outdoors with each other (see Appendix B for more details). Despite this, the mean values of the scenarios clearly demonstrate different epidemiological trends.

**Figure 13:**
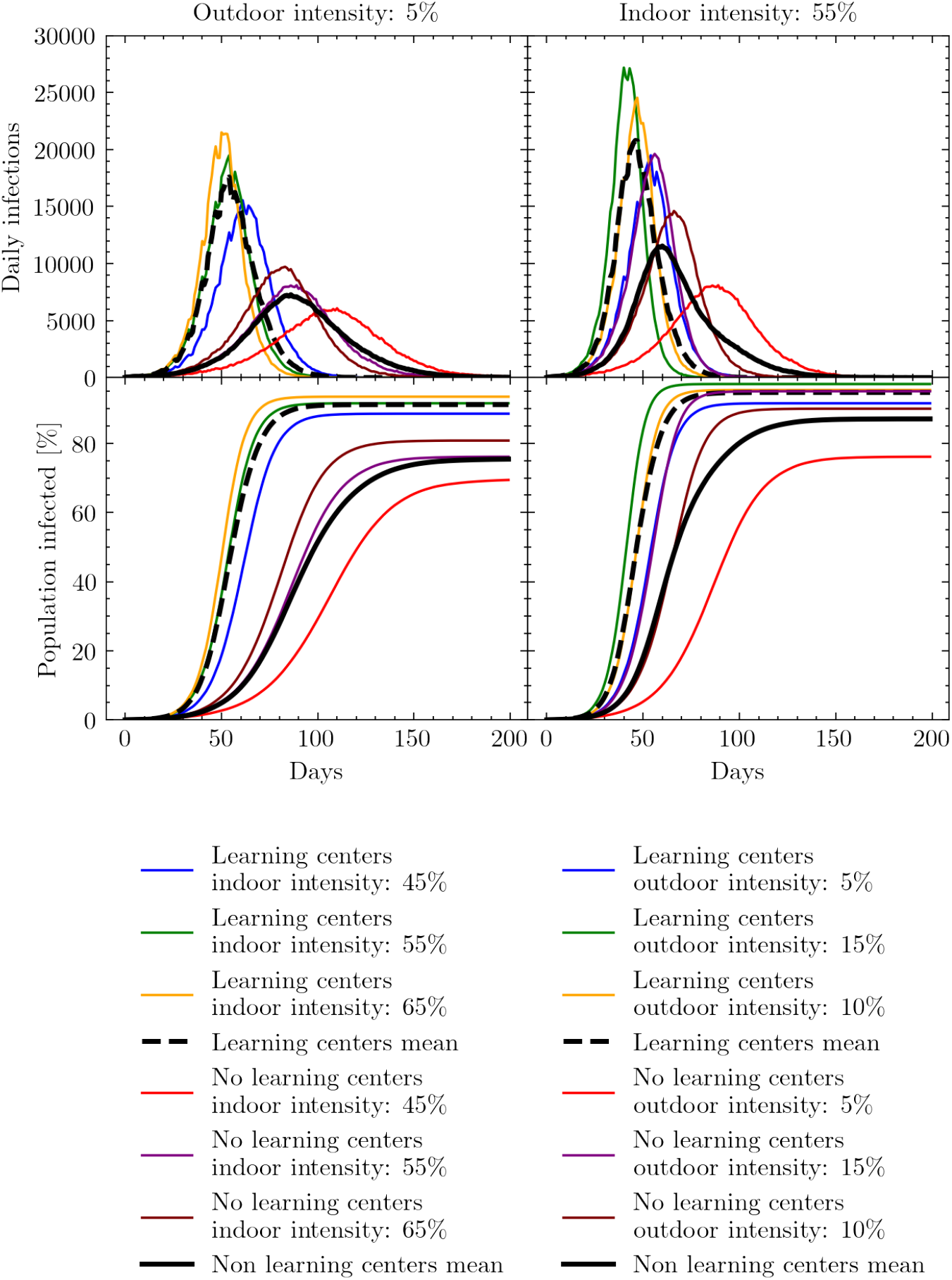
Simulated daily and cumulative infections measured in days since the beginning of the simulation. Results show the effects of varying indoor and outdoor interaction intensity parameters relative to the interaction intensity parameter set for the shelter. These intensity parameters are varied in both the baseline - learning centers remain closed - models and those with learning centers open. Left: the effects of varying the indoor interaction intensity while keeping the outdoor intensity fixed at 5% that of the shelter intensity parameter. Right: the effects of varying the outdoor interaction intensity while keeping the indoor intensity fixed at 55% that of the shelter intensity parameter.

Although opening learning centers may increase both the cumulative number of infections and rate of disease spread, it might be expected that this growth observed in Figure 13 is predominantly constrained to the younger age groups. However, in Figure 14 we see that although opening learning centers does increase the chance of children being infected significantly, this increase in infections rapidly breaks out of age-strata likely due to mixing in often inter-generational shelter settings. This, coupled with the increase in the cumulative number of infections, suggests that by opening learning centers the virus will also likely infect individuals who were previously naturally shielded by a form of herd immunity.

**Figure 14:**
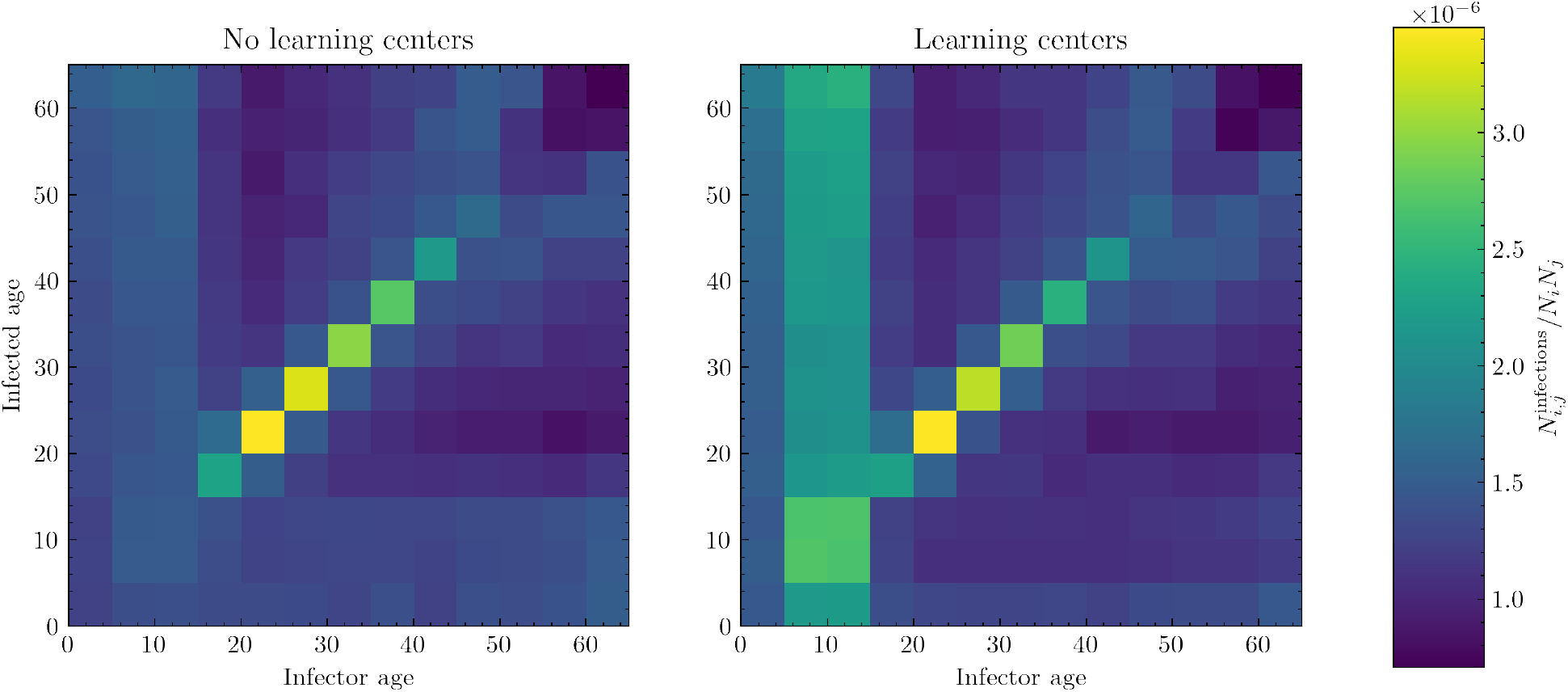
Simulated number of cumulative infections in one age group produced by another age group divided by the size of the infector and the infectee group. Left: the scenario in which learning centers remain closed. Right: the scenario in which learning centers are open. The specific models used to generate these matrices assume the baseline model interaction intensities - i.e. an indoor intensity of 55% that of the shelter intensity and an outdoor intensity of 5% that of the shelter intensity.

### 6.5 Mitigation strategies for opening learning centers

Reopening learning centers is a priority because the longer the learning centers remain closed, the longer children in the settlement go without school and risk having their educational development stunted. In Section 6.4 we found that the opening of learning centers may facilitate the spread of COVID-19. However, the simulations described were only designed to account for the effects of opening learning centers under the same conditions they operated in before the virus was circulating. In this section, we explore several possible strategies for opening learning centers with additional transmission mitigation strategies.

We model three possible strategies: i) adjusting the regularity with which children in the settlement attend learning centers, and therefore limiting their mixing in these environments; ii) opening more learning centers in alternative spaces; and iii) introducing specific measures to lessen the interaction intensities in the learning centers. This final strategy could consist of combinations of physical distancing in classrooms, mask wearing, increasing classroom ventilation, and more thorough cleaning and hygiene.

Since the focus of this section is on proactive policy interventions to mitigate COVID-19 spread in classrooms, we simulate a baseline scenario in which learning centers are closed, and then compare it against alternative scenarios in which learning centers are open with different mitigation strategies. In Section 6.4 we varied our assumptions about indoor and outdoor interaction intensities relative to shelter interaction intensities in order to explore possible learning center transmission scenarios. In this section, we fix indoor and outdoor transmission intensities using the same parameters as in Figure 14, i.e. the indoor intensity (including that in learning centers, absent any policy interventions) is fixed at 55% that of the shelter intensity and the outdoor intensity is fixed at 5% that of the shelter intensity.

The first mitigation strategy we test is changing the regularity with which children attend learning centers. Normally, children enrolled in the educational system are expected to attend their learning center each day; however, by halving the attendance rate and having children only attend once every other day, mixing between different children can be reduced. This intervention would also better enable physical distancing as the number of students in each class is halved. The results of simulating this intervention are presented in the left panels of Figure 15. We see a significant delay in the number of days to peak infection, and a lower percent of the population infected, as a result of alternating the days on which children attend learning centers. While this strategy is clearly effective, we may see additional benefits by combining it with other mitigation strategies.

**Figure 15:**
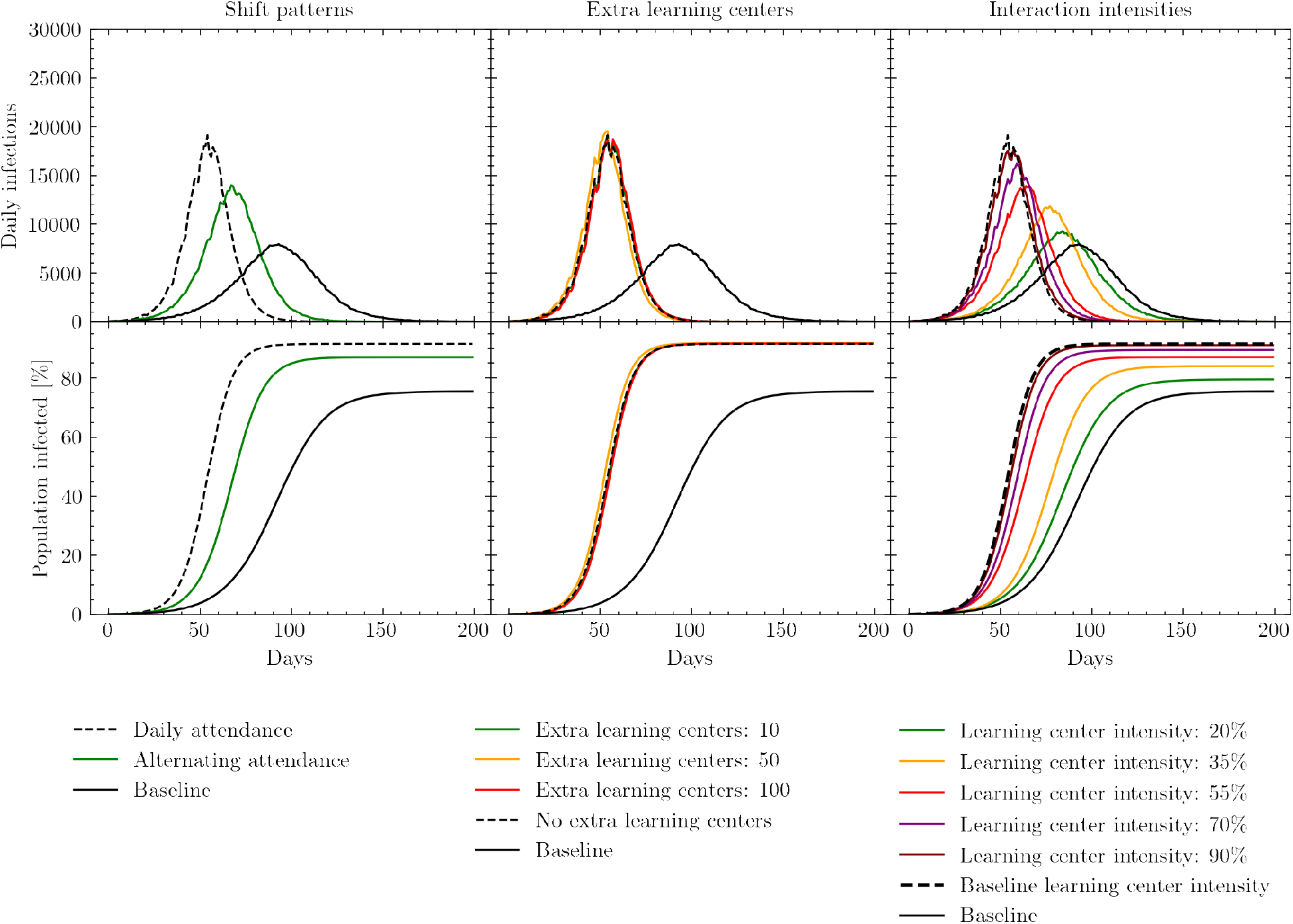
Simulated daily and cumulative infections measured in days since the beginning of the simulation. Black solid lines represent the baseline policy in which learning centers are closed. Black dashed lines represent the policy in which learning centers are open with no additional mitigation strategies. Results show the effects of opening learning centers with three possible transmission mitigation strategies. Left: adjusting the regularity with which children in the settlement attend learning centers. Middle: opening additional learning centers to reduce crowding in the largest classrooms. Right: reducing interaction intensities in the learning centers relative to the baseline learning center interaction intensity, using strategies such as physical distancing, masks, and improved ventilation.

Second, we investigate the possible effects of increasing the number of learning centers in the settlement. To implement this strategy, we first distribute all children to the existing learning centers and then rank the learning centers by those with the biggest class sizes. These large classes pose a higher risk of viral transmission between households, and therefore present a particular danger. Once we have identified the centers with the largest classes, we add another learning center in the same location to our model, thereby mimicking the strategic opening of new learning centers to effectively halve the class sizes of the most crowded centers. Once these new learning centers have been added to the model, we redistribute children from the crowded classrooms to the new centers. As shown in the middle panels of Figure 15, the effect of opening a limited number of additional learning centers is negligible. Relative to the approximately 1,200 learning centers already operating in the settlement, opening 10-100 new centers (which we chose to be the upper-end of a feasible implementation but may already be logistically challenging) does not alter the mixing of children enough across the settlement to have a significant impact.

Finally, we examine the effects of changing the intensity of interactions within the learning centers, while keeping the other indoor, outdoor, and shelter interaction intensity parameters fixed. The range within which we vary these interaction intensities (20-90%) corresponds to the changes in interaction intensities that result from various combinations of physical distancing, mask wearing, and increasing ventilation.

Figure 15 shows the simulated effects of reducing interaction intensity in the learning centers relative to the baseline learning center intensity described above. Reducing learning center interaction intensities has the potential to significantly affect the height and positioning of the daily infection peak, as well as the total number of individuals infected. When learning center interaction intensities fall below 20-35%, daily infection statistics begin to approximate the scenario in which learning centers remain closed, thereby almost completely mitigating the effects of opening the centers. As discussed in Section F.2, the upper end of this relative intensity range could correspond to enforcing mask wearing alone if compliance and the efficacy of the masks worn are high. The lower end may correspond to the combination of physical distancing in classes, mask wearing and improved ventilation [75, 74, 78, 79, 80]. With respect to this last suggestion, it is important to note that ventilation options in schools vary with the type of classroom, e.g. some learning centers are built from bamboo allowing for more natural air to flow, while others appear to be smaller, concrete rooms [81, 82]. In enclosed settings, ventilation could consist of opening windows and doors as well as using electric fans to increase air flow.

In summary, our simulations suggest that implementing a combination of mask wearing, physical distancing, and improved ventilation can significantly decrease the number of infections and potentially make it possible to open learning centers safely. While physical distancing may not be possible in classrooms given the current space available [83], this could be enabled by the reduction of class sizes induced through alternating attendance.

## 7 Data Visualisation Tool

Accurately communicating the detailed data and insights produced by simulations like these is challenging, and many solitary figures eschew detailed plots for a comprehensive overview or vice versa. Another communication challenge arises because correct interpretation of data is often relative: changes in infection numbers only have meaning when we consider factors like the total population, the worst case scenario, and the best case scenario. Policy makers must consider this information in totality. Additionally, inherent uncertainty of hyperparameter values for a given simulation lead to experiments that run simulations over a grid of practicable values, generating a large amount of data that can be difficult to interpret and communicate.

People often turn to dashboards to present interconnected views into large data. However, each of our simulations produces enough information to warrant its own dashboard. Allowing easy comparison of each simulation to another is a significant visualization challenge. To this end, we built an interactive dashboard to accompany our simulations. This tool helps both to explore how the choices of different hyperparameters effect the outcomes of policy decisions and to improve translation between modelers and decision makers.

Our dashboard is built independently of the main simulation code and relies on a detailed data structure collected from an experiment (batch of simulations). Many things can happen to an agent in a single simulation. For example, at any time point an agent can get infected, change locations, infect another, become hospitalized, die, or recover. Each of these ‘events’ can be reduced into a useful statistic (e.g., the number of infected each day or the locations where each agent got infected per day) and stored as a column of a CSV that is eventually consumed by the dashboard along with a metadata file describing the hyperparameters of each run. The dashboard is built with VueJS.^11^ All logic for processing the CSV files and displaying them to the user for interaction lives within the Typescript code. Snippets of primary dashboard functionality are shown in Figures 16, 17, and 18.

**Figure 16:**
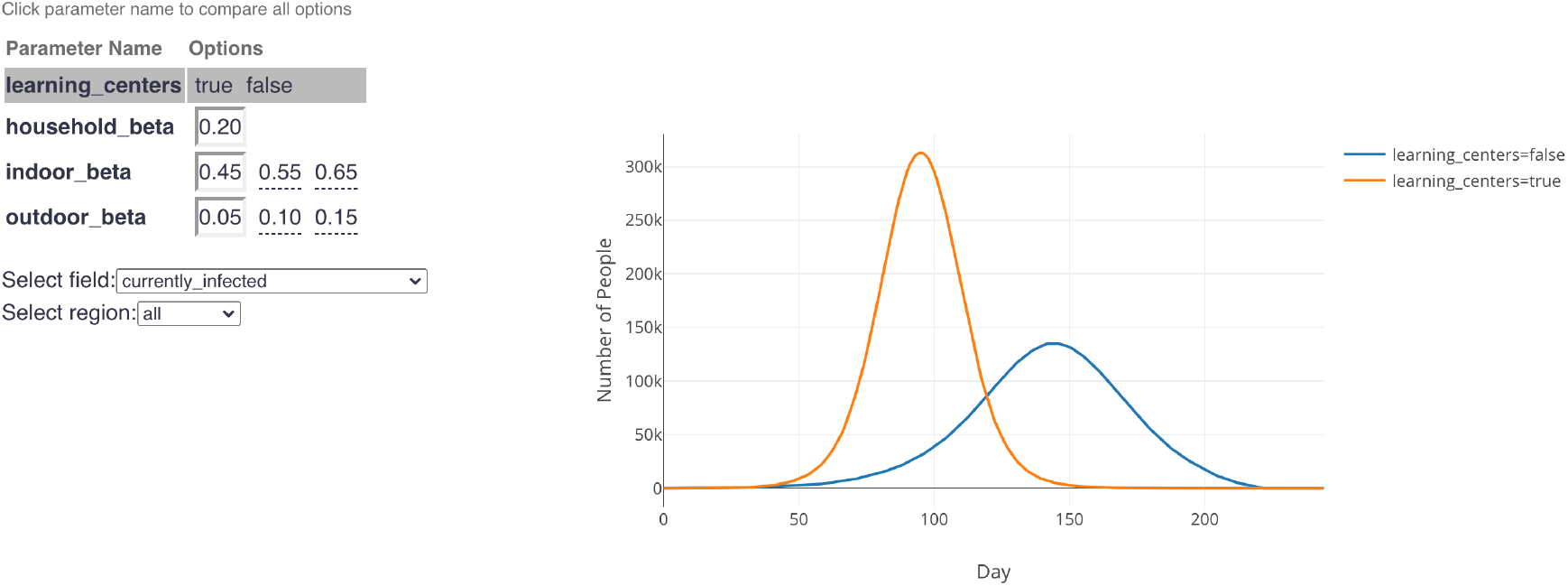
An example from the dashboard showing the infection curves (right) when learning centers are open (orange curve) or closed (blue curve) assuming specified interaction intensities. This graphic answers the question: “How can we expect infection rates to evolve if we open learning centers, assuming people follow mask and social distancing rules such that the indoor infection rate (“indoor beta”) and outdoor infection rate (“outdoor beta”) are low?” Different simulations with different interaction intensities can be selected by clicking different cells in the parameter grid (left). Select the name of a parameter (e.g., “learning centers”, “indoor beta”) to compare the selected field across all different values of that parameter while the other parameters remain constant.

**Figure 17:**
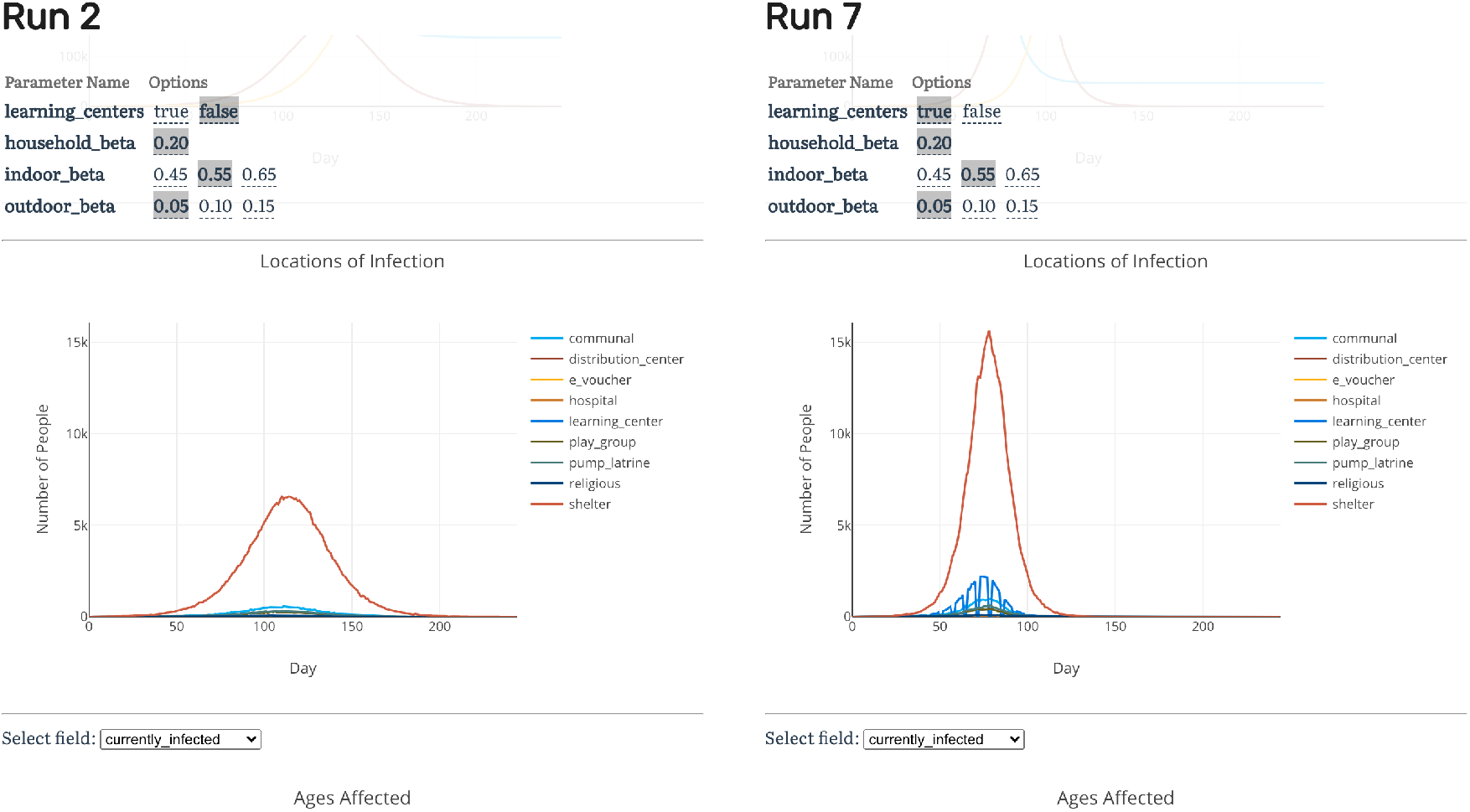
Direct comparison of two different simulations with learning centers open and closed. Displayed plots show distribution of where infections occurred each day (note that infections due to learning centers occur only on weekdays when learning centers are open). Scrolling up or down on this view shows additional comparison plots such as the SIR curve, ages affected, and geographical spread. Select a different run for each column by clicking around the parameter grid (top).

**Figure 18:**
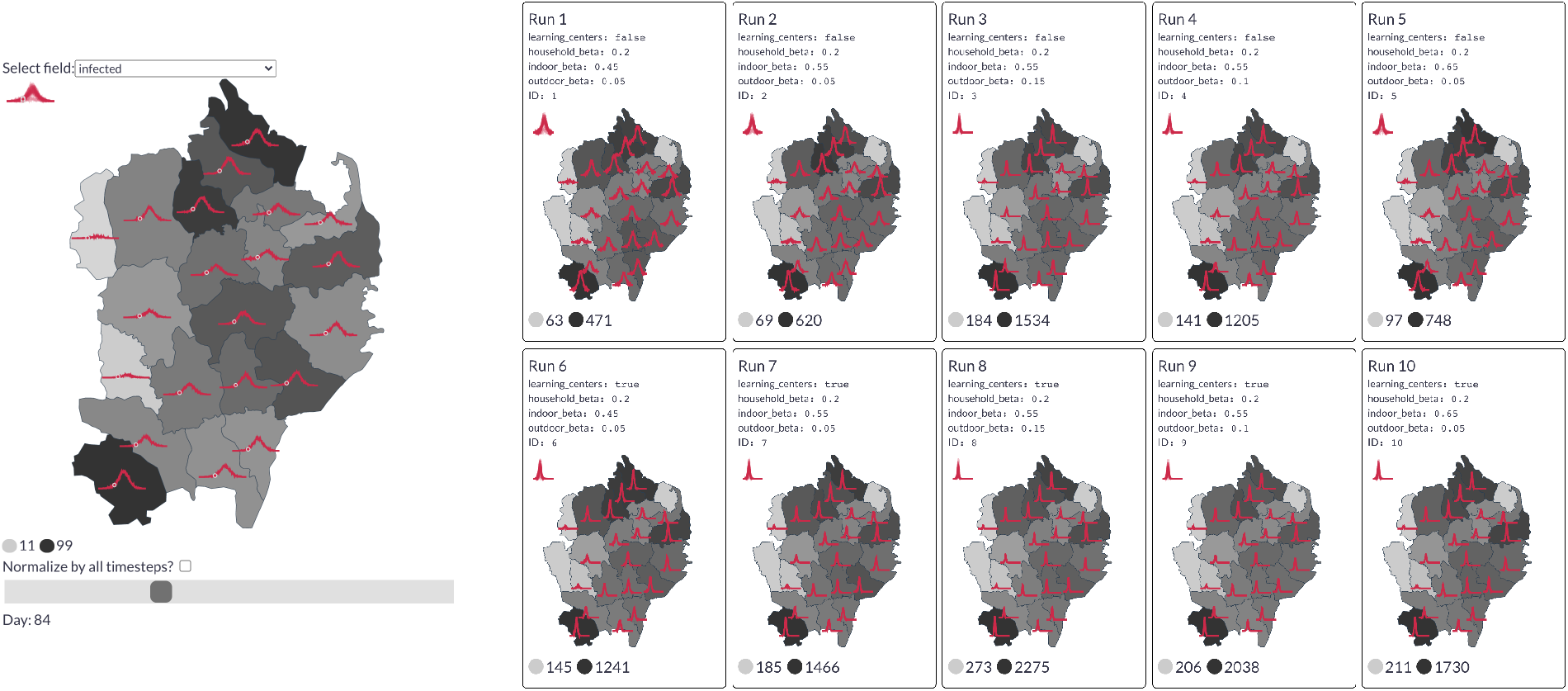
Left: An example from the dashboard showing daily infections by region for a single simulation. The red sparklines indicate the trend over the course of the whole simulation. The circle marks 84 days after the start of simulation (i.e., the current time step). Darker regions indicate a higher infection count at this point in time. Right: Comparison of the daily infection rate of each region across 10 simulations of the “learning centers” project scenario, colored by the maximum peak seen in each region.

The many facets presented by the dashboard assist the discovery of rich insights from the information-dense simulations. In addition to several of the plots shown throughout Section 6, the dashboard allows users to zoom in to simulation results across different parameters for specifics (Figure 17) and zoom out for overviews and easier exploration (Figure 16). It can also show how different interventions vary by geography. How do the initial locations of infections impact the eventual geographical distribution of infections or deaths? How does medical infrastructure play a role in hospitalisation trends? The dashboard plots these trends on top of a regional choropleth map for every simulation and an interactively selected time step. For example, Figure 18 shows that the geographic distribution of infections does not seem to change by simulation, even though the overall height of the infection peaks vary significantly with the chosen parameters.

This dashboard serves primarily as a collaborative tool in three intended ways. For the researcher and data scientist, it enables rapid verification that the collective behavior of the agents aligns with real-world observations. For the policy maker, it provides an extensive view into the potential impact of different policy choices, enabling comparison across different assumptions. Finally, it serves as a communication tool to those unfamiliar with the underlying base model; it exposes the granularity of information that can be extracted from our agent-based model and instills confidence that the ‘best-case’ and ‘worst-case’ scenarios have been sufficiently analyzed.

## 8 Summary

In this paper we present an agent-based modeling approach, adapted from the JUNE framework [10], to simulate disease spread in refugee and IDP settlements. The movement of people and their interactions are modeled at the individual level, with parameters informed by open-source datasets, empirical observations and recent research literature. Our approach first consists of building a ‘digital twin’ of the settlement in which the geographic layout is defined. Virtual individuals are included into the model with different demographic attributes mimicking real world statistics and family and shelter structures are reproduced. Locations in which individuals may interact are also included, such as learning spaces, distribution centers or hand pumps and latrines. Secondly we design a simulation engine which captures what people in the model do during the day, how they interact and how diseases may be transmitted. This underlying structure can then be used to model different operational interventions by altering the movement and interactions of different subsets of individuals in the model, or by closing certain venues. Finally, we present a dashboard designed to present the multiple insights and uncertainties inherent to this modeling approach which can serve as a shared tool for conversation and iteration between modelers and decision makers.

This work focuses on the spread of COVID-19 in the Cox’s Bazar refugee settlement in Bangladesh, although the approach is designed to be generalizable to other settings. Given incomplete testing and case statistics, we have focused on modelling the relative effects of various operational interventions on key statistics such as the daily infection rate, as opposed to producing precise forecasts. Specifically, we analyzed the possible effects of alternate home-case delivery mechanisms, mask wearing based on compliance and the type of masks worn, and (re)opening learning centers under various scenarios. Our findings suggests that the isolation of people with mild to severe symptoms will likely have little effect given the assumed infectiousness profile of symptomatic individuals. Mask wearing, however, is found to be have potentially large positive effects, mitigating significant proportions of disease spread if worn in indoor locations outside shelters. This is the case even if masks such as homemade cotton masks are worn, as opposed to surgical masks. Alternatively, the opening of learning centers could present challenges, with the risk of increasing the growth rate of the epidemic. However, we also explore several strategies to opening learning centers, which if used in combination, could greatly mitigate many of these risks.

One of the main limitations of this work so far has been the possible validation of model predictions with real world data - since case and testing data availability has also been limited. Our approach to understand the potential impact of interventions has been simulating the effects of interventions as if they were in place from the beginning of the simulated period. If required, however, in the event that more precise data becomes available, we expect to be able to perform further retrospective validations of the results by leveraging the flexibility of the model which can be fitted to historical trends, enabling the provision of future forecasts, as well as the simulation of different sequences of measures being implemented at different points in time (see example of this in [10]).

Indeed, a serological study has been carried out throughout December 2020 in the Cox’s Bazar settlement which could serve as a key source of data for fitting, evaluating and constraining our modeling approach. In future work, we also plan to assess the impact of various vaccine distribution strategies in these settings.

In any modeling approach it is important to tailor results and outputs to the specific environments and questions which need to be answered by decision makers. The approach presented in this paper has been developed in close collaboration between modelling teams and those operating in the Cox’s Bazar settlement. Research questions and operational scenarios have been defined jointly by the different teams involved in the project. In fact, we have found that the development of the data visualisation tool plays an important role in helping translate between groups with a wide range of expertise. It is crucial that public health specialist and decision makers have full understanding and ownership of the results of any modeling work. With this work we hope to encourage future multidisciplinary modeling efforts to engage fully with end users to ensure meaningful discussions take place and decisions are taken informed by the best possible science.

## Data Availability

In the interest of openness and transparency, the simulation code has been released under a GPL v3 license. Given the sensitivity of this situation, more information about the datasets used and how to access them is available upon request.

https://github.com/UNGlobalPulse/UNGP-settlement-modelling

## Acknowledgements

United Nations Global Pulse work is supported by the Government of Sweden, and the William and Flora Hewlett Foundation. JB, CCL, AQB and AS are also supported by the UKRI-STFC grant number ST/P006744/1. The UK Public Health Rapid Support Team is funded by UK aid from the Department of Health and Social Care and is jointly run by Public Health England and the London School of Hygiene Tropical Medicine. The University of Oxford and King's College London are academic partners. The views expressed in this publication are those of the authors and not necessarily those of the National Health Service, the National Institute for Health Research or the Department of Health and Social Care. We would like to thank the whole JUNE team for their assistance in adapting this models. We would like to thank the whole JUNE team for their assistance in adapting this models. We would especially like to thank Frank Krauss from Durham University and Kevin Fong from University College London for their support and guidance with this project. We would like to thank Leonardo Milano from the United Nations Office for the Coordination of Humanitarian Affairs (OCHA), Ahmed El Saeed and Patricia Loh from the United Nations Technology Innovation Labs (UNTIL) Finland, and Keyrellous Adib from the WHO East Mediterranean Regional Office for their comments and support. We would like to thank the GridPP team at Durham University and Manchester University for their support and computing time spent on their systems. This work used the DiRAC@Durham facility managed by the Institute for Computational Cosmology on behalf of the STFC DiRAC HPC Facility (www.dirac.ac.uk). The equipment was funded by BEIS capital funding via STFC capital grants ST/K00042X/1, ST/P002293/1, ST/R002371/1 and ST/S002502/1, Durham University and STFC operations grant ST/R000832/1. DiRAC is part of the UK’s National e-Infrastructure.

This paper made use of Python [84] and the following Python libraries: Matplotlib [85], Numpy [86], Pandas [87], Scipy [88], SciencePlots [89]. This paper also made use of GNU parallel 2018 [90].

## Ethics

This research has been designed and conducted following relevant data privacy and data protection principles and processes, including UNHCR data protection policies and guidelines, as well as the UN principles on Personal Data Protection and Privacy, and the UNSDG Guidance Note on Big Data for the 2030 Agenda: Ethics, Privacy and Data Protection. Data used for building the digital twin come from statistical data, other open datasets and anonymous and aggregated survey data used collected by UN agencies as cited throughout this document.

## Code availability

In the interest of openness and transparency, the simulation code has been released under a GPL v3 license and can be accessed from: https://github.com/UNGlobalPulse/UNGP-settlement-modelling. More information about the datasets used and how to access them is available upon request.

## A Parameters

In this section we detail the data sources for both the digital twin and simulation parameter ranges. By specifying the parameter choices made in the context of the Cox’s Bazar settlement, we hope to enable simple adaptation of this approach to other settings, given appropriate available data.

### A.1 Digital twin

Table 2 specifies the data sources for parameters relating to specific inputs for the creation of the digital twin of the Cox’s Bazar settlement. These inputs include the locations of different infrastructure and facilities; population demographics and household composition; and access to health care as well as data on comorbidities.

**Table 2:** Data sources for the digital twin.

| Description | Source |
| --- | --- |
| Population Demographics and Geography | UNHCR & Government of Bangladesh [28], IOM [42] |
| Household Composition | UNHCR & Government of Bangladesh [28], UNHCR [5] |
| Access to Healthcare | UNHCR [12] |
| Pump & Latrine Locations | UNHCR [5] |
| Infrastructure Locations | ISCG [56] |
| School Attendance & Children’s Activities | Cox’s Bazar Education Sector [50], Oxfam [51], GAGE/IPA/YALE [52, 53] |
| Additional Hospital Locations | ISCG [91], Truelove et al. [92] |
| Comorbidity Data | Clark et al. [43], Global Burden of Disease study [46], UN Population Estimates [93] |

### A.2 Simulation

Table 3 specifies the data sources for parameters relating to specific inputs used in the creation of the simulator, which models the movement of people in the Cox’s Bazar settlement. These inputs include information on the frequency of visits to facilities such as markets, distribution centers, and religious centers, dis-aggregated by age and sex.

**Table 3:** Data sources for the simulator including literature relating to how often individuals attend different locations based on demographic characteristics.

| Description | Source |
| --- | --- |
| Distribution Centers | Hoddinott [55], Oxfam [51], ISCG & Government of Bangladesh [94, 95], UNHCR [96], WFP [70, 69] |
| Non-Food Distribution Centers | ISCG [56] |
| E-Voucher Outlets | WFP [69, 70, 97], Hoddinott [55] |
| Communal Centers (kitchens, centers, etc.) | ACAPS [7], Government of Bangladesh [67], WV [98], ISCG [56] |
| Female-friendly Spaces | UN Women [99], WFP [100] |
| Market Attendance | AAH [101] |
| Religious Center Attendance | Lopez et al. [102] |
| Learning Centers | HRW [54], Government of Bangladesh [68], Cox’s Bazar Education Sector [50], ISCG [56], WFP [70] |
| Pump & Latrine Usage | UNHCR [5, 103], WFP [70] |

## B Activities

In the previous section we detailed the data sources for both the digital twin of the Cox’s Bazar settlement and the simulator to model the movement of people. In this section we discuss how these sources and others are used to derive assumptions which guide our choice of model inputs. Table 4 contains information regarding each of the possible activities which individuals in the model may perform at different time steps. We specify details of individual movements during normal operations (i.e. pre-COVID-19) and how these movements have changed since March 2020 when the settlement began introducing measures to mitigate the spread of COVID-19. Details explicitly included in our model are highlighted with an asterisk.

**Table 4:**
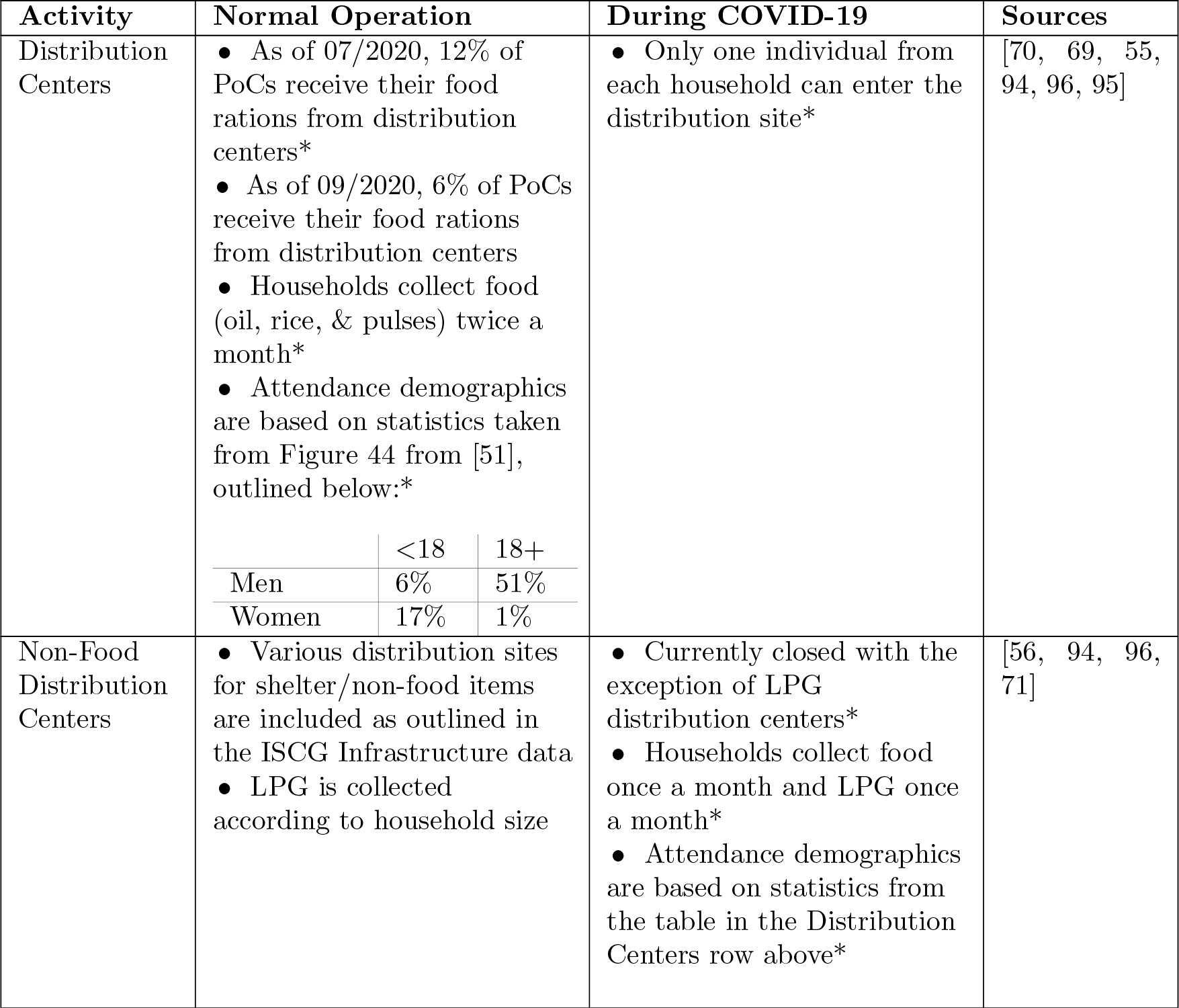
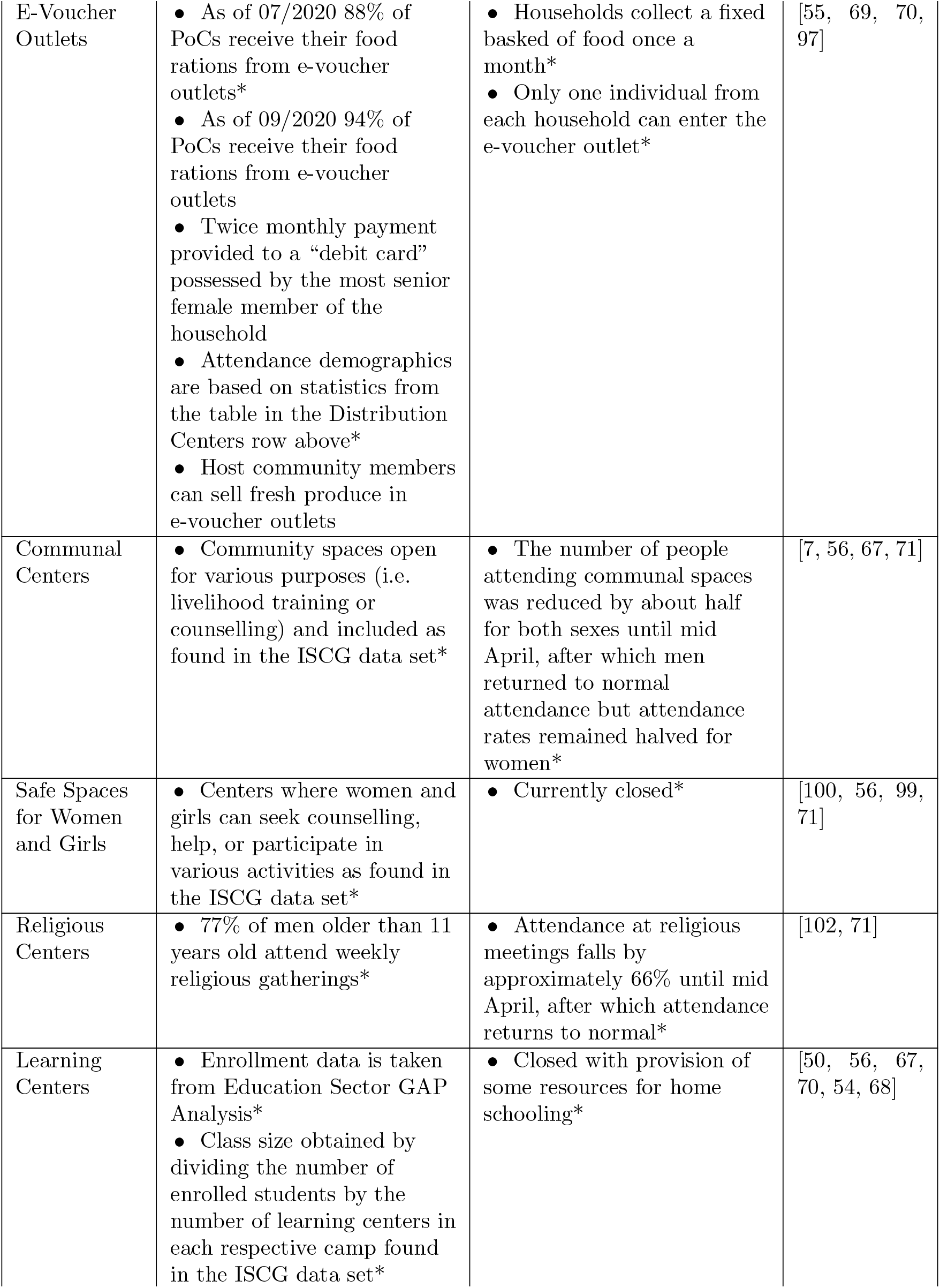
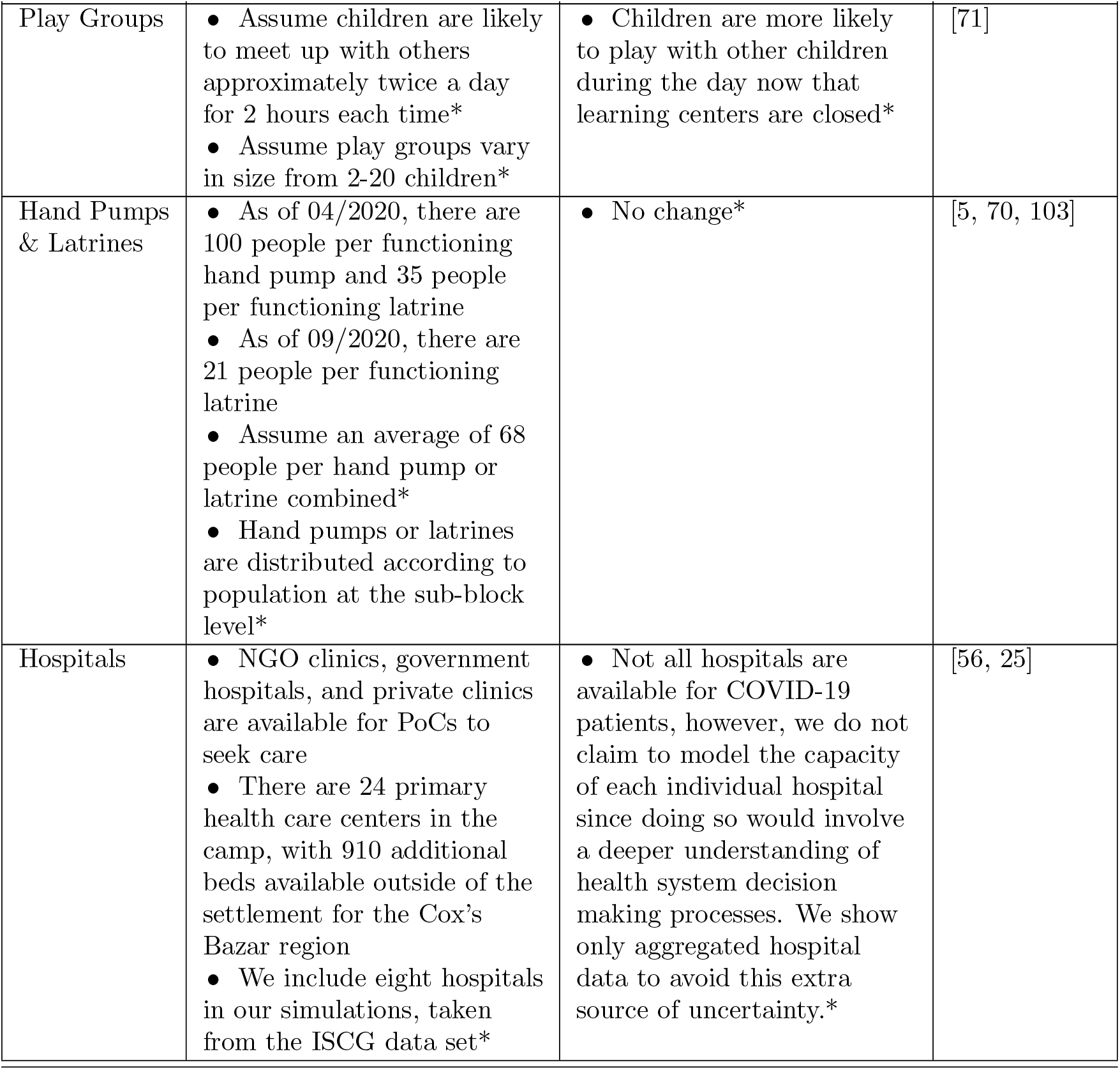
Details and assumptions used in each activity included in the simulation. * indicates that the specific detail is currently being implemented in the simulation.

## C Mixing Matrices

Contact matrices are used by many modeling approaches, including ABMs, to specify the number of contacts between different people. Common sources of these matrices are surveys such as [58, 59, 60] from which contact matrices can be derived stratified by age and type of location in which the contact was made. Many of these surveys also delineate between physical and conversational contacts, the former being more intense than the latter. As mentioned in Section 5.2, we account for these different types of contact through specifying a physical contact multiplier which represents the proportion of all contacts in a given setting which are physical in nature.

While many surveys exist at the national level, it is not clear how these apply in the settlement setting in which living conditions, and cultural norms, can vary greatly from their host country, or of the PoC’s country of origin. However, a key advantage of the JUNE framework [10] is that the precise contact matrices do not need to be fully known in order to reproduce the age and location dependent mixing patterns found in reality. This is because, in reality, contact matrices are largely a function of an individual’s movement patterns and the activities they participate in. Since we naturally incorporate this information in the setting of age and sex dependent probabilities that an individual goes to a certain location, then we just need to focus on getting the broad character of the contacts correct, e.g. encouraging school children to be more likely to interact with each other than the teacher in a learning center. Furthermore, given the form of Equation 5.5, the scaling of the *β*^(*L*)^(*t*) parameter by location *L*, the setting of which we discuss in detail in Appendix D.2, absorbs the uncertainty of the total number of contacts in each location. A similar logic applies to the proportion of contacts which are physical in nature. A full discussion of this can be found in Section 4 and Appendix B of [10].

Adopting the naming convention of [10], we will call the input to our model “social mixing matrices” and the derived output the “contact matrices”. Here we discuss the input social mixing matrices for each location which are designed to capture the broad character of the interactions in these venues. Given the degree to which full contact matrices are unknown in this setting, we aim to keep the input social mixing matrices simple so as to capture reasonable mixing but not overly bias results.

The choices of values in this section are chosen such that all matrices have a similar order of magnitude effect with the exception of learning centers, where the asymmetry in contacts between members of the participant groups (teachers and students) is most pronounced. This means that when we group indoor and outdoor intensity parameters (*β*^(*L*)^ parameters) together for scenario modelling, as in Section 6, we can create new scenarios easily by altering the relative values of these parameters without having to account for the multiplicative effect of the social mixing matrices. Furthermore, since this paper focuses on modeling the relative efficacies of interventions, some uncertainty in the relative social mixing matrix values is further divided.

### C.1 Shelters

Some shelters in the Cox’s Bazar settlement, and other settlements, are shared between up to two families. We assume that within each family, all individuals are equally likely to interact with each other given the proximity of the setting. If a family shares a shelter with another family then we assume that each family is half as likely to interact with the other family than within themselves. For the proportion of physical contacts, we assume this scaling to be a factor of 4 different with 80% of contacts within a household being physical, but only 20% of those with another household who shares the same shelter. This results in:

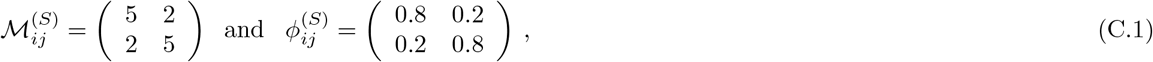

where {*i, j*} ϵ {*H, O*} for the household and other household entries respectively. Here, 5 is chosen as the intra-household contact baseline in order to ensure all social mixing matrices have a similar order of magnitude effect on the probability of transmission.

### C.2 Learning Centers

Learning centers have two key groups which can interact, students, *S*, and teachers, *T*. To ensure that students and teachers make proportionately the correct number of contacts given the average student-teacher ratio of 30:1 we set:

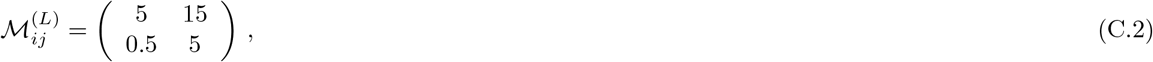

where {*i, j*} ∈ {*T, S*} which is in close agreement with that derived in Appendix B.2 in [10].

We assume the proportion of physical interactions to be the same among all participants in the learning center given the closeness of the classroom setting and so this is absorbed in the intensity parameter value.

### C.3 Play groups

When modelling interaction in play groups, for simplicity we assume children in the same age bracket play and interact with each other and do not interact with others outside their age bracket. We define age brackets to be: 3-6 year olds, *P*1, 7-11 year olds, *P*2, and 12-16 year olds, *P*3. The proportion of physical interactions are assumed to scale by age bracket:

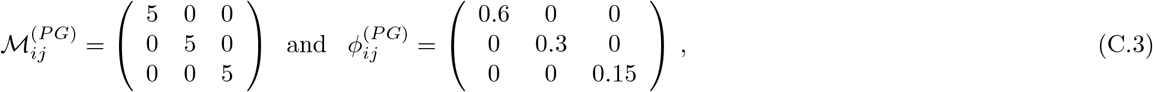

where *{i, j} ∈ {P* 1, *P* 2, *P* 3*}*.

### C.4 Other locations

Given the lack of available data on contact patterns, in all other locations in which interactions can occur, e.g. distribution centers and communal centers, we assume the social mixing and physical contact matrices to be single valued at 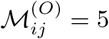 and 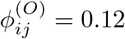 respectively. This follows the same logic as that in Appendix B.4 of [10].

## D Seeding and intensity parameters

### D.1 Initial seeding

To initialize the infection we use data from the WHO EWARS datasets [13] on confirmed cases by camp on the 24th of May. By that time, only six of the camps had detected at least one case of COVID-19, with case numbers totaling 22 across the Expansion Site. To account for under-reporting and other factors we conservatively scale this number by a factor of four. To seed the infection, we assume 88 people are COVID-positive and infectious at the beginning of the simulation. Half of these case are assumed to be distributed in the six camps with confirmed cases weighted by their relative prevalence, and the other 44 cases are randomly distributed among the remaining camps relative to their population size. This choice means that after the first day of simulation time, all results presented in Section 6 have surpassed 100 cases.

### D.2 Intensity parameter selection

The interaction intensity parameters (*β*^(*L*)^ parameters - see Equation 5.5) control multiplicatively the intensity of interactions in different locations *L*. In the event where historical data is available to fit to, thereby enabling model predictions, these parameters would act as free fitting parameters [10]. However, in settings with limited available data on COVID-19 cases and other statistics, fitting a model to data is not possible. To account for this we focus on modeling the relative effects of different operational interventions, rather than precise statistics, and set these intensity parameters heuristically.

As mentioned in Section 6.1, we group together intensity parameters into three categories: shelter, indoor and outdoor. Each location, and its corresponding intensity parameter is categorised into one of these groups following the location type given in Table 1. While in reality each of the locations in a given group might have a different intensity parameter value, for simplicity we will assume the members of each group share the same intensity parameter value unless specified otherwise, i.e. people interact in all indoor locations with the same intensity with the exception of in the shelter environment, and people interact in all outdoor locations with the same intensity.

To set the parameters we first assume a hierarchy of values: shelter *>* indoor *>* outdoor. We then assume that the interaction intensity in indoor spaces outside the shelter environment is approximately half that of inside the shelter. This choice is designed to heuristically capture the fact that you will be less likely to interact with others outside your family in non-shelter settings as intensely as you would interact with your own family in the shelter. Given the density of shelters in the Cox’s Bazar settlement, we feel this is a reasonable assumption. Although, there is little research on the subject, what studies have been carried out suggest that outdoor transmission is significantly less likely than indoor [64, 65, 66]. Indeed, reports making explicit calculations comparing the probability of transmission estimate that indoor transmission could be as much as 18-19 times more likely than outdoor [65]. Since we cannot give a definitive quantification of this intensity, we fix this parameter conservatively at 5% of the shelter interaction intensity parameter.

In some scenarios, since we are modeling the relative effects of interventions, the exact choices of these assumed values are less important. However, as shown in Section 6, when modelling scenarios in which the specific interaction intensity ratios become more dominant, we vary the ratios of the intensity parameters to explicitly capture some of this uncertainty.

In all scenarios the value of the shelter intensity parameter is fixed and all other intensity parameters varied relative to this value. To arrive at a value for the shelter intensity we scanned several possible values while fixing the relative indoor and outdoor intensity values as stated above. Figure 19 shows the results of this parameter scan over a 200 days period since the beginning of the simulation. Early data from the WHO EWARS dataset [13] did not suggest a large initial surge of infections in the camp of the magnitude as might be expected by high value choices of the shelter intensity parameters. Therefore we heuristically choose a value of 0.25 since this scenario allows for different scenarios of disease transmission, both more and less severe than this baseline thereby enabling easy comparison. From the perspective of the timing of the peak, this scenario is in good agreement with the ‘moderate transmission’ scenario presented in [25].

**Figure 19:**
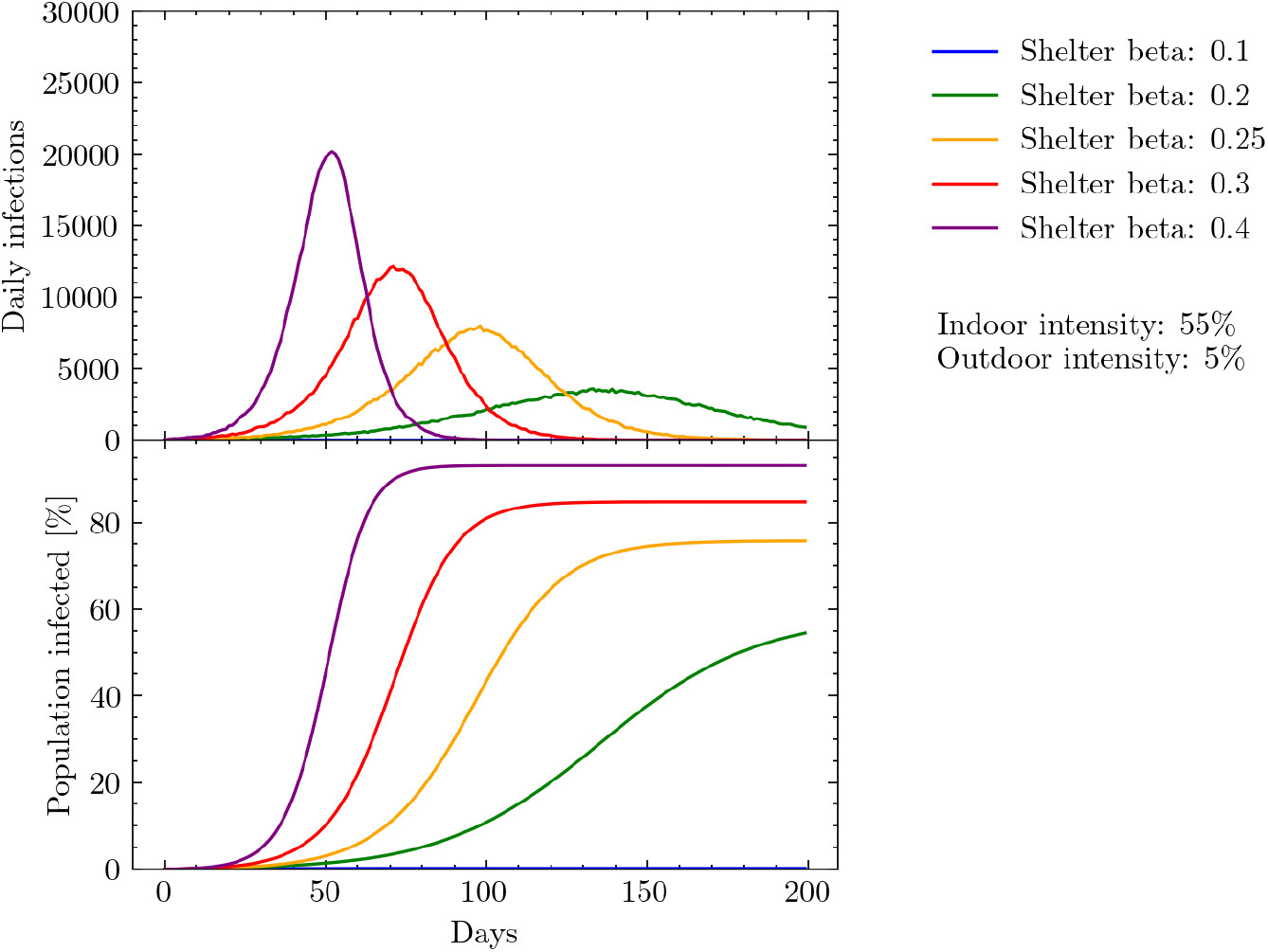
Simulated daily and cumulative infections measured in days since the beginning of the simulation. Results show the effects of varying the shelter intensity parameter while keeping the indoor and outdoor intensity parameters fixed at 55% and 5% respectively relative to the shelter intensity value.

It should be noted that since the value of the shelter intensity parameter remains fixed in all scenarios, and no scenario modeled thus far has altered this intensity parameter, that the parameter simply acts as a scaling factor for the growth rate of the disease across all scenarios. Therefore, its relative effects, again, will largely divide out when comparing across scenarios making scenario-based modeling less sensitive to this parameter value. Furthermore, Figure 19 clearly demonstrates our model’s ability to simulate a wide range of different transmission scenarios if required, along with its flexibilty to fit to historical data.

## E Comorbidities

Relative to previous simulations in refugee settings, one of our primary contributions is the inclusion of population-specific comorbidities in our model of COVID-19 risk. Accounting for such comorbidities is important because much of the data on COVID-19 severity comes from the developed world, where the prevalence of chronic illnesses such as diabetes or cardiovascular disease may be unusually high, and the prevalence of infectious diseases such as HIV/AIDS and tuberculosis may be relatively low. When using developed-country estimates of COVID-19 risk to estimate risk in the developing world, the differing prevalence of chronic illnesses may lead to an upward bias in the estimated risk of severe illness due to COVID-19, whereas the differing prevalence of infectious diseases may lead to a downward bias in the estimated risk of severe illness due to COVID-19. Therefore, properly estimating location-specific severity rates requires accounting for these differences in comorbidity distributions.

Our approach to accounting for comorbidities is based on the work of Clark et al. [43], who provide estimates of how 12 different types of comorbidities influence the relative risk of developing severe disease as defined by “severe acute respiratory illness (fever and at least one sign/symptom of respiratory disease, e.g. cough, shortness of breath; AND requiring hospitalization)”.12 The complete list of conditions and risk multipliers is included in Table 5. Given detailed estimates of the risk of developing severe COVID-19 by age and sex in one country, Clark et al. develop a risk adjustment method to modify these estimates for the comorbidity mix of another country, which we adapt for our purposes and describe in more detail below.

**Table 5:** Comorbidity multipliers taken from Clark *et al*. [48]. These multipliers represent the relative risk of developing severe COVID-19, conditional on being affected by the comorbidity in question. A healthy individual has a relative risk of 1, implying that e.g. an HIV-positive person has a 1.5-times greater risk of developing severe COVID-19 than a healthy person.

| Risk Group | Risk Score |
| --- | --- |
| HIV/AIDS | 1.5 |
| Tuberculosis | 1.5 |
| Cancers with direct immunosuppression | 1.5 |
| Cancers with possible immunosuppression | 1.5 |
| Cardiovascular diseases | 3 |
| Chronic respiratory diseases | 1.5 |
| Diabetes mellitus | 3 |
| Chronic kidney diseases | 3 |
| Chronic neurological disorders | 1.5 |
| Sickle cell disorders | 1.5 |

We begin with data on the average risk of developing severe COVID-19 conditional on infection by age and sex cohort in the UK.^13^ Presumably, these risk estimates already take into account the comorbidities which affect the relevant cohort in the UK. Therefore, we divide these average risk estimates by an adjustment factor in order to ‘purge’ the UK statistic of specific attributes relevant only to the UK population. The result is effectively a comorbidity-free probability of developing severe COVID-19 conditional on age and sex.

Concretely, we calculate the comorbidity-free age- and sex–specific risk of infection as:

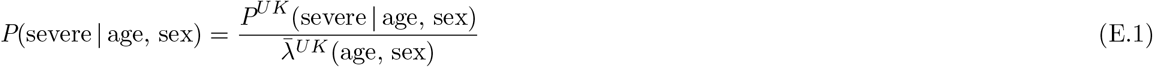

where *P*^*UK*^(severe | age, sex) is the probability of developing severe COVID-19 for a given age and sex cohort in the UK as described above and *λ*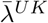(age, sex) is the UK-specific adjustment factor. The latter is defined as:

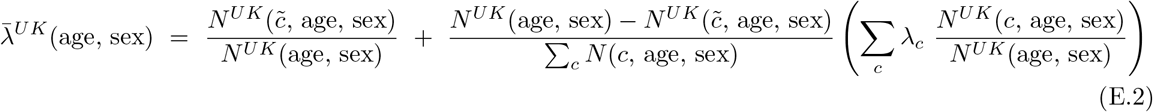

where 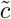 denotes individuals with no comorbidities;^14^ *c* indexes comorbidities; *λ*_*c*_ is the corresponding comorbidity multiplier from Table 5; *N* ^*UK*^(*c*, age, sex) represents the number of people in a given age and sex cohort that are afflicted by comorbidity *c* in the UK; and *N* ^*UK*^(age, sex) represents the total number of people in the age and sex cohort in the UK. The term in parentheses in Equation E.2 is a weighted sum of comorbidity multipliers, where the weights represent the fraction of the population with each comorbidity in the UK. The prefactor that multiplies the term in parentheses is a normalization term to ensure that the population fractions across all comorbidity conditions and the no-comorbidity condition sum to 1. This is necessary because a single individual may have multiple comorbidities, so the sum of empirically-observed population fractions over all comorbidity conditions could be greater than one.

We next attach a calculated age-, sex- and comorbidity-specific risk of severe infection to each agent in the model. Recall that agents are drawn from a multivariate age, sex, and comorbidity distribution. The distribution of age and sex in the population is derived from settlement statistics (see Appendix A), whereas the distribution of comorbidities is drawn from the age- and sex-specific comorbidity prevalence in Myanmar as estimated by the 2017 Global Burden of Disease (GBD) study and tabulated in Clark *et al*. [46, 47].^15^ For each sampled individual, we calculate the risk of severe disease by adjusting the comorbidity-free risk estimate by the multiplier from Table 5 that is specific to the individual’s assigned comorbidity:

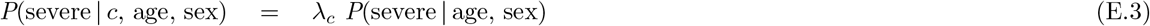

This probability of severe infection can then be used to simulate the trajectory of each individual’s illness, if infected.

## F Intervention Details

### F.1 Isolation centers

This simulation compares a scenario in which symptomatic patients go to isolation and treatment centers if they have mild or severe symptoms, but do not require hospitalization, to a scenario in which patients with mild and severe symptoms self-quarantine and are treated at home (referred to as ‘home-based care’). The time delay between the onset of symptoms and going to a treatment center is drawn from a Normal distribution. The mean of this distribution is varied across simulations between 0-5 days but the standard deviation was fixed at 1 day. We also vary the time spent in the isolation and treatment center before being released between 5-10 days, and the compliance rates with which symptomatic individuals present themselves for testing between 50-100%.

In reality, tests used in the Cox’s Bazar settlement take approximately 2 days to be processed and therefore choosing a mean time delay to isolation of 2 days presents a reasonable scenario. Including time delays as low as 0 days with high compliance rates presents a best-case scenario in which tests can be rapidly processed.

### F.2 Mask Wearing

The effect of mask wearing in different locations depends on both mask efficacy and mask wearing compliance. Much of the literature on the efficacy of mask wearing is in preliminary stages and/or has small sample sizes, making it difficult to draw precise conclusions from the results. In addition, several studies focus on exploring the percentage of droplets which pass through masks, rather than on calculating the effect of masks on reducing the probability of transmitting the virus, since the latter is often difficult to infer and can be impacted by many additional factors such as the distance between people wearing masks and whether masks are worn by both parties in an interaction or just by one [78].

Despite these caveats, the efficacy of masks can be estimated based on mask filtration ability, mask fit, and mask reuse [104]. Mask filtration depends on the number of layers of fabric and the type of fabric used. Masks with a greater number of layers can be more effective at filtering but might be less comfortable, particularly in humid environments like Cox’s Bazar, which could potentially reduce compliance [78]. Research also suggests that mask fit has a large influence on efficacy, as improper fit (large gaps between the face and mask) can result in up to a 60% decrease in filtration capability [105].

As a result of these factors, varying assumptions about mask efficacy and compliance are useful for estimating the true effects of mask wearing. We define compliance as the rate of people wearing masks correctly, whereas efficacy is defined as a function of the mask material, as well as any degradation through incorrect reuse and washing. For example, assuming no large gaps (i.e. mouth and nose are covered during use), along with other proper use and no degradation, we can assume mask efficacy for single-layer cotton and surgical masks to be approximately 50% and 80% respectively [77, 74, 78]. However, these efficacies may be reduced due to reusing or washing them [106, 104]. Indeed, these is a common problem in the settlement where mask supplies are limited and in the early phases of the epidemic, many PoCs were having to wash and reuse surgical masks not designed for this [71].

To account for these possible scenarios, we varied the compliance rate with correct mask wearing between 10-100% and the efficacy of the masks between 10-90%.

## Footnotes

1 ‘Persons of Concern’ is a general term that encompasses populations of interest to UNHCR with varying legal statuses, including “refugees, returnees, stateless people, the internally displaced, and asylum-seekers” [2].

2 Time reported using an average 2010s processor and requires at least 1 gigabyte of computer memory to run.

3 We note that a national level characterization could be biased, however, there is currently a lack of available data at the relevant demographic scale. People living in remote locations with various predisposing risk factors may not be able to be generalized at the national level and vise versa. The use of comorbidity data is only used in our model to adjust the likelihood of disease progression and therefore the chance that someone may be hospitalised. Since we take the approach of assessing the relative efficacies of operational interventions these uncertainties will often be sub-dominant, however, in the event of any precise predictions these uncertainties should be clearly stated.

4 We use the term ‘household’ interchangeably with ‘family’, however, the former can often be more accurate in settings where individuals might live together but are not technically from the same family.

5 A survey is underway to better constrain the time spent by individuals in different locations to inform future modeling efforts. However, for now we will make the assumption of four to five 2-hour time slots per day. It should be noted that the precise time spent in a given location is fixed for simplicity as it allows for the direct comparison of contact matrices in these locations (see Equation 5.5).

6 This choice of nearest locations is decided heuristically and can be set by location-type as required if data is available to constrain this.

7 Currently, Poisson parameters are determined only based on an individual’s age and sex, however this can be adapted to other demographic and behaviour variables if data is available.

8 We are in the process of collating data from a recent survey carried out in the Cox’s Bazar settlement to help inform the derivation of such mixing matrices.

9 Note that the *α*(*t*^*!*^) parameter serves an important purpose when fitting parameters to historical data, although this is not the focus of this work.

10 While data on healthcare seeking behaviour in the Cox’s Bazar settlement is incomplete, there is a strong community based referral system which could be a source of data for future modeling efforts.

11 https://v3.vuejs.org/

12 Note that in our model, we allow for the possibility of severe cases and death without hospitalization.

13 This is the setting in which the JUNE model was developed, and was therefore an already-available source of detailed COVID-19 data.

14 Comorbidity prevalence rates are provided for each comorbidity, and we do not have data on how often particular comorbidities coincide. Therefore, we do not know how many people have no comorbidities. To calculate this number we follow the general strategy outlined in Clark *et al*. [43]. First, we assume that comorbidities are independently distributed, and calculate the expected proportion of people with one or more comorbidities; we then scale this proportion by 0.9 to account for likely correlations between comorbidities. Next, we confirm that the estimated proportion of people with one or more comorbidities is greater than the proportion of people with each single comorbidity; otherwise, we set the estimated proportion of people with one or more comorbidities equal to the proportion of people with the most common comorbidity. Subtracting this number from 1, we obtain the estimated proportion of people with no comorbidities. That is, 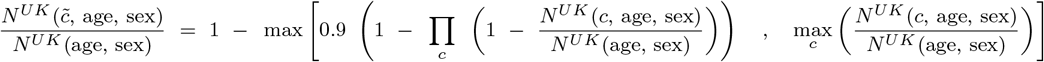

15 While there are active efforts to track illness in the camps, these efforts are oriented towards communicable disease and did not provide sufficient data to estimate the prevalence of the diseases in Table 5. Therefore, we have used the population of Myanmar as the reference population for comorbidity prevalence estimates.

